# The Cystic Fibrosis Transmembrane Regulator Controls Tolerogenic Responses to Food Allergens in Mice and Humans

**DOI:** 10.1101/2024.06.17.24309019

**Authors:** Marc Emmenegger, Chryssa Zografou, Yile Dai, Laura R. Hoyt, Ravindra Gudneppanavar, Andra Chincisan, Hubert Rehrauer, Falko J. Noé, Natalia Zajac, Georg Meisl, Matthias M. Schneider, Hang Nguyen, Katja Höpker, Tuomas P. J. Knowles, Mireia Sospedra, Roland Martin, Aaron M. Ring, Stephanie Leeds, Stephanie C. Eisenbarth, Marie E. Egan, Emanuela M. Bruscia, Adriano Aguzzi

## Abstract

IgE antibodies against the allergen Ara-h2 can cause life-threatening anaphylaxis upon exposure to peanuts. Desensitization strategies aim at inducing IgG responses against Ara-h2 which may compete with anaphylactogenic IgE. Here we assessed anti-Ara-h2 titers in an unselected cohort of 24,536 adult patients admitted to a general hospital for disparate medical reasons. Surprisingly, adult (n=177) and pediatric (n=76) patients with cystic fibrosis (pwCF) had IgG4, but not IgE, against several peanut and soybean allergens, yet did not suffer from peanut allergy. Antibody repertoires were not globally perturbed in pwCF, and heterozygous Cystic Fibrosis Transmembrane Regulator (*CFTR*) mutation carriers had the same prevalence of food allergies as pwCF. Peanut sensitisation of *Cftr^-/-^* mice failed to induce IgE and was associated with elevated IFN-γ. We conclude that CFTR is a key regulator of anaphylactogenic and tolerogenic responses to food allergens. CFTR-controlled cytokine responses including IFN-γ, in combination with a compromised epithelial barrier, may trigger a preferential IgG4 response resulting in tolerance to food allergens.

**Highlights:**

- We investigated serum IgG against Ara-h2 in 24,536 patients and identified 133 seropositives.
- Seropositivity was associated with cystic fibrosis, and these patients had IgG4 but not IgE.
- We reproduced these results in a paediatric validation cohort and in a *Cftr^-/-^* mouse model.
- A compromised epithelial barrier and the cytokine milieu might explain this phenotype.

## Introduction

Considerable quantities of food-derived antigens are absorbed through the intestinal epithelia every day. This requires oral immunological tolerance to these intrinsically innocuous food constituents. Oral tolerance permits selective responsiveness to food antigens or commensals, a phenomenon conventionally termed ‘systemic unresponsiveness’ (Moran & Burks, 2015; Pabst & Mowat, 2012; Tordesillas & Berin, 2018). Mucosal immunoglobulins (Ig) A directed against food constituents are common in non-allergic individuals (Berin, 2012; Frossard et al., 2004; E. G. Liu et al., 2022); serum IgG to some food allergens is occasionally detected in those without food allergy (Husby et al., 1985; Leviatan et al., 2022), and antigen-experienced CD4^+^ T cells are required for the induction of tolerance (Lockhart et al., 2023). Conversely, malfunctioning oral tolerance is associated with IgE-mediated hypersensitivity reactions to food allergens whose prevalence in industrialized countries has sharply risen over the past thirty years (Branum & Lukacs, 2009; Perkin et al., 2016), as has the prevalence of other chronic diseases (Agache et al., 2022; C. A. Akdis, 2021; C. A. Akdis & Nadeau, 2022). Food allergies entail a high level of morbidity and contribute to mortality worldwide. While different therapeutic interventions have been tested and applied (Durham & Shamji, 2022), many food-related allergic diseases remain incurable (Wood et al., 2024).

Passive immunotherapy with monoclonal IgG antibodies that bind the allergen and prevent IgE engagement are promising candidates to alleviate the disease burden (Orengo et al., 2018; Paolucci et al., 2023; Shamji et al., 2021) and may become an important instrument in treating anaphylactogenic allergies, like peanut allergy (PA). Although naturally occurring mucosal antigen-specific IgA may not predict tolerance (E. G. Liu et al., 2022), systemic blood-based antibody signatures, assessed in sizeable populations, can reveal immunological mechanisms of natural tolerance and highlight the prevalence of antigen-specific antibodies in certain diseases. Conversely, they can point to their association with absence of disease. We have investigated the antibody repertoires of large collections of patients for the occurrence of autoantibodies against the prion protein (Senatore et al., 2020), the tau microtubule-binding domain (Magalhães et al., 2021), and apoptosis-associated speck-like protein containing a CARD (Losa et al., 2023), and have studied seroepidemiological correlations of immune reactivity to SARS-CoV-2 proteins in 72,250 plasma samples of 54,153 individuals (Emmenegger, De Cecco, et al., 2023).

Here, we have interrogated a large unselected hospital cohort for the presence of IgG against the Arachis hypogaea 2 (Ara-h2) protein, the major peanut allergen (Fig**. 1A**, overview). We identified 127 seropositive among 24,536 patients, of which about 0.2% (n=45) suffered from documented peanut allergy. An exploratory data-driven analysis indicated a strong association with cystic fibrosis (CF), a life-limiting autosomal recessive monogenic disorder prevalent in Europe, North America, and Australia (Elborn, 2016; Rowe et al., 2005). We propose that the combination of gut epithelial dysregulation and with the inflammatory milieu typical of persons with CF (pwCF) may trigger enhanced antigen penetration, here Ara-h2, and may elicit an IgG4 dominated B cell response.

**Fig. 1.**
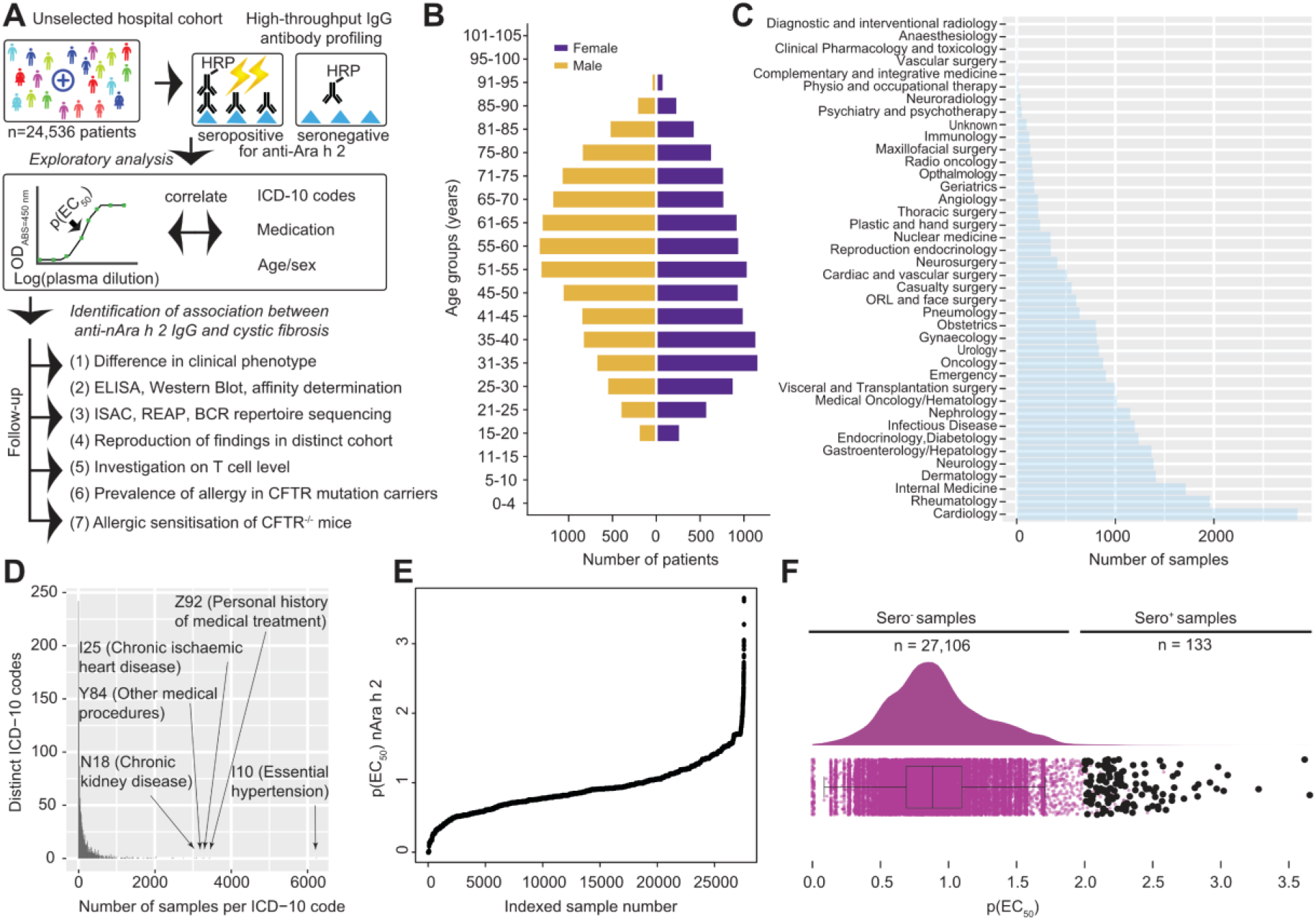
High-throughput serological screening for IgG antibodies against the natural Ara-h2 (nAra-h2) allergen in an unselected university hospital cohort. **A**. High-throughput serological screen (HTS) and follow-up conducted in this study. **B**. Age/sex pyramid of patients included in the HTS. **C.** Clinical departments in charge of patients whose samples were incorporated in the HTS. **D.** Histogram of ICD-10 codes ascribed to patients included in this study. Arrows: most frequently assigned ICD-10 codes. **E.** Index plot with p(EC_50_) values for all patients included in the screen. **F**. Dots: individual p(EC_50_) values of all samples. Distributional information: half-violin plot. Black dots: seropositive samples.

## Results

### High-throughput antibody profiling identifies individuals with IgG against Ara-h2

Using an automated high-throughput screening (HTS) platform (**Fig. 1A**), we assayed the presence of IgG antibodies directed against “natural” (i.e. purified from delipidated extract by multi-step chromatography) Ara-h2 (nAra-h2) in 24,536 patients admitted to the University Hospital Zurich (USZ) for a broad variety of reasons. The plasma samples (n=27,239) had been collected for routine diagnostics and patients had consented to their use for biomedical research. Our cohort comprised 48% female (n=11,944) and 52% male (n=12,592) patients with a median age of 54 (IQR: 49-68) years (**Fig. 1B** and **Table 1**). No selection was imposed on this cohort other than being treated as inpatients or outpatients in any of 39 clinical divisions of the University Hospital of Zurich.

**Table 1.** Population characteristics for different analyses performed in this study.

| Assay | Characteristics | Non-hits | Hits | Total |  |
| --- | --- | --- | --- | --- | --- |
| *High-throughput screening* | Individuals, number | 24,409 | 127 | 24,536 |  |
|  | Median age (IQR), years | 54 (49-68) | 45 (32-58.5) | 54 (49-68) |  |
|  | Sex, female % | 48 | 42 | 48 |  |
|  | Sex, male % | 52 | 58 | 52 |  |
| Assay | Characteristics | Non-hits | Hits | Total |  |
| *CF profiling* | Individuals, number | 88 | 31 | 119 |  |
|  | Median age (IQR), years | 33 (25-40) | 31.5 (25.25-37.25) | 32 (25-40) |  |
|  | Sex, female % | 49 | 46 | 47 |  |
|  | Sex, male % | 51 | 54 | 53 |  |
| Assay | Characteristics | CF | PCF | PC | HC |
| *Secondary screen (ELISA)* | Individuals, number | 177 | 76 | 319 | 358 |
|  | Median age (IQR), years | 34 (26-42) | 13 (9-17) | 54 (39-67) | 44 (31-55) |
|  | Sex, female % | 51 | 45 | 48 | 60 |
|  | Sex, male % | 49 | 55 | 52 | 40 |

The largest collections of samples originated from the Departments of Cardiology, followed by Rheumatology, Internal Medicine, Gastroenterology and Hepatology, Dermatology, and Neurology (**Fig. 1C**). Accordingly, the spectrum of diseases was highly heterogeneous and encompassed 1,532 individual ICD-10 three-letter codes. The five most frequent ICD-10 codes observed were I10 (‘Essential (primary) hypertension’, n=6,235), Z92 (‘Personal history of medical treatment’, n=3,437), I25 (‘Chronic ischaemic heart disease’, n=3,282), Y84 (‘Other medical procedures as the cause of abnormal reaction of the patient, or of later complication, without mention of misadventure at the time of the procedure’, n=3,237), and N18 (‘Chronic kidney disease’, n=3,095), as annotated in **Fig. 1D**.

Approximately 25% of individuals (n=6,280) had some indication of having self-anamnestic, suspected, confirmed, or non-confirmed allergy, and 0.2% (n=45) reported peanut allergy, which reflects the approximately expected frequency of clinically diagnosed peanut allergy in adult European populations (Kotz et al., 2011; Nwaru et al., 2014; Spolidoro et al., 2023). Other common allergies and food intolerances had the following roughly estimated prevalence in the dataset: nut allergy without further specification (0.4%, n=103), lactose intolerance (0.7%, n=163), fish allergy (0.2%, n=44), pollen allergy (1.5%, n=377), house dust mite allergy (0.6%, n=155), cat hair allergy (0.4%, n=103), dog hair allergy (0.1%, n=19), penicillin allergy (4.9%, n=1197), paracetamol allergy (0.5%, n=126), and nickel allergy (0.8%, n=190). The most frequently prescribed medications (from a total n=5,854 medications listed) were paracetamol (n=11,409), metamizole (n=8,992), esomeprazole (n=6,338), pantoprazole (n=5,874), and cholecalciferol (n=5,254). The sampling for the screen occurred during a period from August 2016 to December 2019 and the primary HTS was conducted over the same time span with a break between March 2018 and October 2019.

For serological analyses, plasma samples were serially diluted using a twofold geometric progression (range: 1:50-1:6,000) in high-protein binding 1536-well plates using contactless acoustic dispensing. Optical densities (ODs) were recorded as reported (Emmenegger, De Cecco, et al., 2023) and logistic regression curves were derived. Antibody titers were defined as the inverse decadic logarithm of the dilution at the inflection point of the sigmoid (-log_10_(EC_50_), abbreviated as p(EC_50_)) **(Fig 1E**). p(EC_50_) values (≥ 1.7) aligned well with the averaged ODs at the three highest concentrations (Pearson correlation coefficient R = 0.90, **Fig. S1A)**. We visualized individual data in half-violin plots (**Fig 1F**). Seropositive samples were defined as having p(EC_50_) values ≥ 2, corresponding to half-maximal binding at a 1:100 plasma dilution with mean squared residual error of the logistic regression < 20% of the p(EC_50_) value (black dots in **Fig. 1F**) based on cut-offs previously defined (Senatore et al., 2020). Values < 2 were regarded as seronegative. Thus, we detected 133 (0.49%) seropositive samples from 127 seropositive individuals.

### Association between antibodies against nAra-h2 and Cystic Fibrosis

Seropositive samples showed a balanced proportion of females (n=53) and males (n=74), similarly to seronegative samples (χ^2^, p = 0.238, **Fig. 2A**). The median age of seropositives (45, IQR: 32-58.5 years) was significantly lower than that of seronegatives (54, IQR: 49-68 years; Mood’s median test for age: p < 0.00001, **Fig. 2B**). The largest fraction of the seropositive patients came from Pulmonology/Pneumonology, Dermatology, or Gastroenterology and Hepatology, with a five-fold increase seen for Pulmonology (p < 0.0001, Fisher’s exact test, **Fig. 2C**). None of the other clinical divisions displayed a statistically significant enrichment of seropositive patients.

**Fig. 2.**
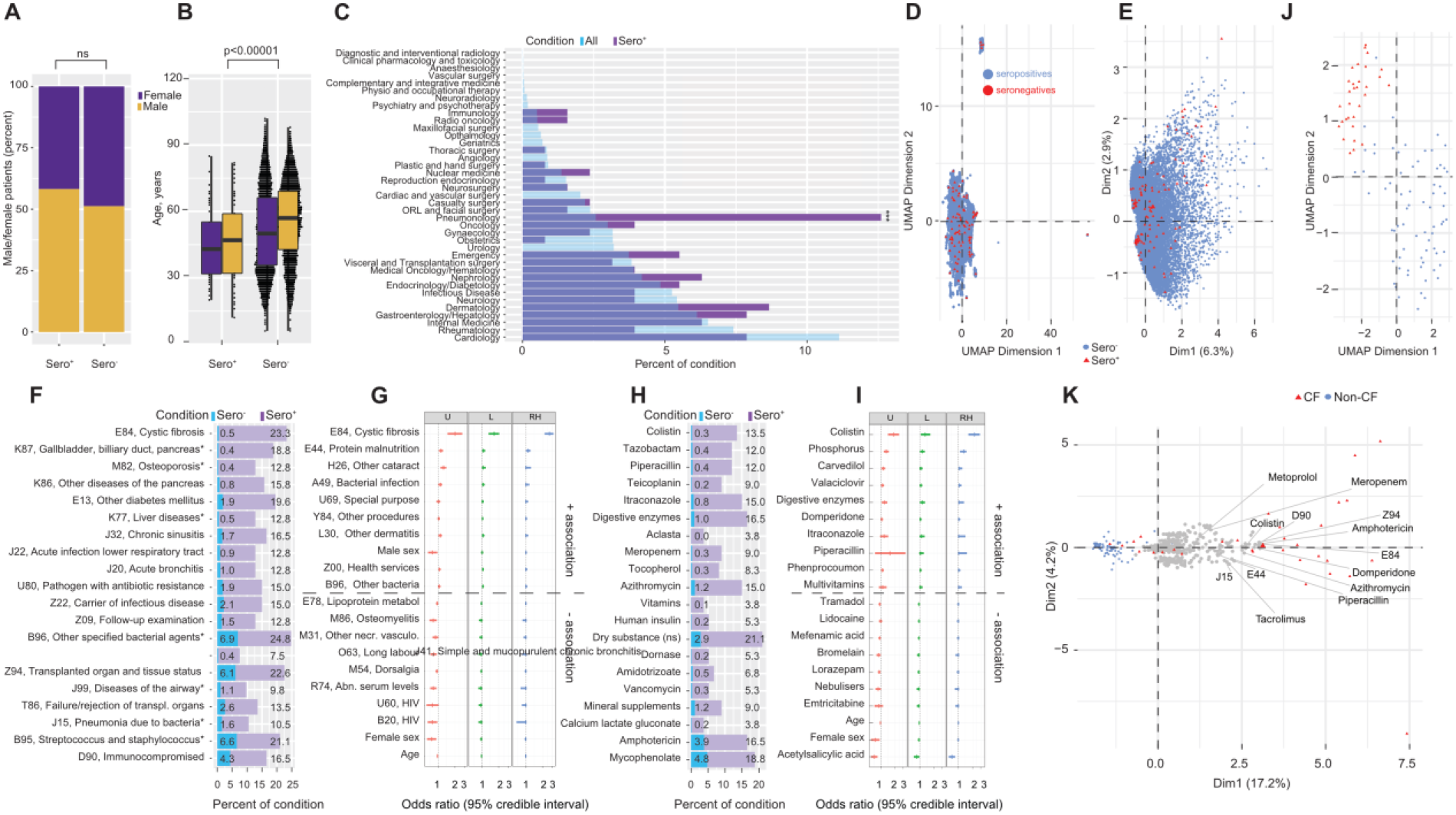
Association between IgG to Ara-h2, CF, and CF comorbidities. **A**. Sex distribution in Ara-h2 seropositive and seronegative individuals. Ns: Χ^2^ non-significant. **B**. Age of Ara-h2 seropositive and seronegative females and males. Mood’s median test for age was significant (p < 0.00001). **C.** Contributions of clinical departments to seropositive individuals (%). Enrichment of seropositives for pneumonology (p < 0.0001, Fisher’s exact test). **D-E.** UMAP (D) and PCA (E) analysis of ICD-10 codes and sex showing similar clusters of seropositive and negative fractions. **F**. Top 20 enriched ICD-10 codes, all statistically significant (χ^2^ p < 0.0001 adjusted for multiple comparisons). **G**. Multiple logistic regression analysis using Bayesian priors (U: unregularized. L: LASSO. RH: Regularised horseshoe) showing the ten top positive and top negative odds ratios for ICD-10 codes, age, and sex. **H**. Top 20 significantly enriched medications (χ^2^, p < 0.00001 adjusted for multiple comparisons). **I**. Multiple logistic regression analysis using Bayesian priors (U: unregularized. L: LASSO. RH: Regularised horseshoe) showing the ten top positive and negative odds ratios for medication, age, and sex. **J**. UMAP analysis using all ICD-10 codes, medication and sex for all seropositive individuals. **K**. PCA of the data in (J) with annotation of important features.

We then asked whether the seropositive patients possessed distinguishing features on a global scale. We employed Uniform Manifold Approximation and Projection (UMAP) and Principal Component Analysis (PCA) to project the maximum variability contained in the dataset, containing 1,532 ICD-10 codes and sex information from 23,035 unique individuals and 25,597 samples. Neither UMAP (**Fig. 2D**), using a cosine metric to account for binary data, nor PCA (**Fig. 2E**) revealed any markedly distinct clusters between seropositive and seronegative samples.

To better understand the features characteristic of the seropositive samples, we collected ICD-10 codes occurring at least 5 times (i.e. in > 3.75%) in the seropositive group and conducted χ^2^ statistical testing (*p* = 0.01) applying Bonferroni’s correction for multiple comparisons. We obtained a table of 131 ICD-10 codes, 36 of which displayed statistical significance (**Supplemental File 01**). The most enriched ICD-10 code, E84 (Bonferroni-corrected p < 10^-10^), encodes CF, the most common autosomal recessive monogenic disorder in the Caucasian population. CF was followed by ‘diseases of the gallbladder, biliary tract, and of the pancreas, with diseases elsewhere classified’ (K87, Bonferroni-corrected p < 10^-10^), ‘osteoporosis, with disease elsewhere classified’ (M82, Bonferroni-corrected p < 10^-10^), ‘other diseases of the pancreas’ (K86, Bonferroni-corrected p < 10^-10^), ‘other form of diabetes mellitus’ (K77, Bonferroni-corrected p < 10^-10^), ‘chronic sinusitis’ (J32, Bonferroni-corrected p < 10^-10^), and ‘acute infection of the lower respiratory tract’ (J22, Bonferroni-corrected p < 10^-10^), among others (**Fig. 2F**). As many of these codes appeared to be related to CF, we removed patients with ICD-10 code E84 from the dataset and repeated the analysis. Although some ICD-10 codes fulfilled the criteria of being shared by ≥ 5 seropositive individuals, only one of the ICD-10 codes reached statistical significance (L70, ‘Acne’, see **Supplemental File 02**) whereas all previously significantly enriched codes disappeared. This confirms that ICD code E84 dominated the dataset, with multiple other ICD-10 codes co-segregating with CF. Indeed, diabetes (Gibson-Corley et al., 2016; Kayani et al., 2018), osteoporosis (Gibbens et al., 1988; Hecker & Aris, 2004), chronic sinusitis (Davidson et al., 1995; Okafor et al., 2020; Ramsey & Richardson, 1992) and infections (Cosgriff et al., 2020; Lyczak et al., 2002) are known highly prevalent comorbidities of CF (Bell et al., 2020; Cutting, 2015; Elborn, 2016) expected to be enriched within this patient group.

The threshold of p(EC_50_) ≥ 2 is arbitrary and results in binarization of the dataset (seropositive and seronegative patients). We therefore repeated the analysis on a continuous scale, using the entire spectrum of p(EC_50_) values (**Fig. 1F** and **Fig. S1B** for qq-plot). We conducted a regression on logit-transformed EC_50_ values (denominated as -logit(EC_50_)) in a Bayesian framework using an uninformative (normal(0,10) and two shrinkage priors: LASSO (Gelman et al., 2013) and a regularized horseshoe (Piironen & Vehtari, 2017). We then plotted the odds ratio and the 95% credible intervals of the 10 top-most positively and negatively associated features (**Fig. 2G**). The ICD-10 code for CF (E84) was the only parameter unambiguously correlating with high -logit(EC_50_) values, corroborating results presented above (odds ratio with horseshoe prior: 2.66, 95% credible interval: 2.18-3.14). E44 (“Protein-energy malnutrition of moderate and mild degree”) displayed a slightly increased odds ratio, followed by ICD-10 codes H26 (“Other cataract”), and A49 (“Bacterial infection of unspecified site”). Among the ICD-10 codes that were found to be most negatively associated are HIV-related codes B20 (“Human immunodeficiency virus [HIV] disease resulting in infectious and parasitic diseases”) and U60 (“Clinical category of HIV disease”). Female sex and age were slightly negatively associated with -logit(EC_50_) values, but the decrease in odds ratio was minimal (with horseshoe prior: 0.99, 95% credible interval: 0.97-1.00) and the associations are most likely spurious. Odds ratios for all ICD-10 codes are listed in **Supplemental File 03**.

Next, we used the same χ^2^-based approach to correlate the active principles of medication prescribed to the patients. Of 114 active ingredients appearing at least 5 times in the seropositive fraction, 45 were statistically significant after Bonferroni correction (**Supplemental File 04**). The following medications were associated with Bonferroni-corrected p p < 10^-10^: Colistin (last-resort antibiotic medication for multidrug-resistant Gram-negative bacteria), Tazobactam, Piperacillin, Meropenem and Teicoplanin (against Gram-positive bacteria including methicillin-resistant Staphylococcus aureus), antifungal compounds like Itraconazol and Amphotericin, digestive enzymes, and Tacrolimus (immunosuppressant, p < 10^-6^) (**Fig. 2H**). All these drugs are typically used for the treatment of CF, and indeed the removal from the dataset of pwCF led again to the disappearance of any significantly enriched medication. The application of afore Bayesian multiple regression model identified candidates similar to those found with χ^2^ statistics (**Fig. 2I**) including Colistin (odds ratio with horseshoe prior: 2.19, 95% credible interval: 1.63-2.87), phosphates, Carvedilol and Piperacillin. The HIV medication Emtricitabine was among the most negatively associated features, in line with ICD-10 codes U60 and B20 (**Fig. 2G**, **Supplemental File 05)**. As a limitation, the regression model – unlike the χ^2^ analysis – was restricted to active ingredients assigned to at least 100 patients in the data set to increase the computational robustness.

The seropositive collective (n=127) with p(EC_50_) ≥ 2 consisted of 31 pwCF and 89 with other diseases but no record of CF. Seven patients for whom no information was available were excluded from the analysis. Using ICD-10 codes, active ingredients, and sex as features, we clustered the seropositives, with ICD-10 code E84 as classifier. UMAP and PCA (**Fig. 2J-K**) revealed a distinct cluster formed by pwCF with characteristic ICD-10 codes (e.g. D90, E44, E84, J15, Z94) and medications (e.g. Meropenem, Amphotericin, Domperidone, Piperacillin, Colistin) aligning along the axis of the first principal component (**Fig. 2K)**.

In summary, we found an association between IgG antibodies against the major peanut allergen nAra-h2 and CF, but not with any other disease (**Table 2)**. Similar studies performed with the cellular prion protein, PrP^c^ (Senatore et al., 2020) and with calcitonin-gene related peptide (CGRP), in part with a largely overlapping collection of samples, did not indicate any association with CF. The seropositives did not overlap with PrP^c^ or CGRP reactivity (**Fig. S1C**) except for one individual seropositive for both nAra-h2 and PrP^c^. Prior reports do not suggest a higher prevalence of peanut allergy or other food allergies in pwCF (Lucarelli et al., 1994), and additional data on allergies indicated that only two of the seropositive pwCF had an anamnestically reported peanut allergy (1.6%) whereas penicillin allergy was reported in twelve seropositive individuals (9.5%).

**Table 2.** Number and prevalence of select gastrointestinal, autoimmune, and inflammatory diseases recorded in individuals included in the primary HTS. The prevalence was calculated by dividing the number of occurrences of patients with the ICD-10 code by the total number of patients screened.

| Disease | ICD-10 | Number of occurrences | Prevalence in % |
| --- | --- | --- | --- |
| *Cystic fibrosis* | *E84* | 169 | 0.6 |
| *Gastritis and duodenitis* | *K29* | 906 | 3.3 |
| *Functional dyspepsia* | *K30* | 88 | 0.3 |
| *Crohn's disease* | *K50* | 217 | 0.8 |
| *Ulcerative colitis* | *K51* | 220 | 0.8 |
| *Other and unspecified noninfective gastroenteritis and colitis* | *K52* | 541 | 2.0 |
| *Irritable bowel syndrome* | *K58* | 113 | 0.4 |
| *Intestinal malabsorption* | *K90* | 102 | 0.4 |
| *Systemic lupus erythematosus* | *M32* | 181 | 0.7 |
| *Connective tissue disease* | *M35* | 594 | 2.2 |
| *Type I diabetes mellitus* | *E10* | 437 | 1.6 |
| *Type II diabetes mellitus* | *E11* | *2440* | *9.0* |
| *Asthma* | *J45* | *504* | *1.9* |
| *Other chronic obstructive pulmonary disease* | *J44* | *949* | *3.5* |
| *Rheumatoid arthritis* | *M05 and M06* | *798* | *2.9* |
| *Other disorders involving the immune mechanism* | *D89* | *43* | *0.2* |

### Post hoc clinical profiling does not distinguish between seropositive and seronegative pwCF

First, we investigated whether ICD-10 codes of pwCF with antibodies against nAra-h2 (n=31) differ from those without (n=88). Neither hierarchical clustering (**Fig. 3A**), PCA (**Fig. 3B**), or UMAP (**Fig. 3C**) detected differences in the assigned codes. Also age (p: 0.8217, Fisher’s exact test) and sex (p: 0.7187, Wilcoxon rank sum test) were equally distributed in pwCF with and without anti-Ara-h2 antibodies (**Fig. 3D**-**E** and **Table 1**). The seropositive and seronegative pwCF did not significantly differ in CRP as well as leukocyte and lymphocyte counts (Wilcoxon rank sum test with Benjamini-Hochberg correction, **Fig. 3F**) suggesting that disease exacerbation was no decisive factor. Because forced expiratory volumes in the first second FEV1 scores – an important feature of lung function – were not available in our dataset, we could therefore not classify lung disease severity. We therefore investigated selected features of CF pathology (Bruscia & Bonfield, 2016, 2022; De Lisle & Borowitz, 2013; Dorsey & Gonska, 2017; Elborn, 2016; Garg & Ooi, 2017; Nichols & Chmiel, 2015; Ooi & Durie, 2016; Roesch et al., 2018). The distributions of Ara-h2 antibody-positive and negative pwCF did not differ regarding the presence of a lung transplant (p = 0.6185, Fisher’s exact test), infection with *Pseudomonas aeruginosa* (p = 0.412, Fisher’s exact test), pancreatic insufficiency (p = 0.5727, Fisher’s exact test), or aspergillosis (p = 0.7994, Fisher’s exact test, **Fig. 3G**).

**Fig. 3.**
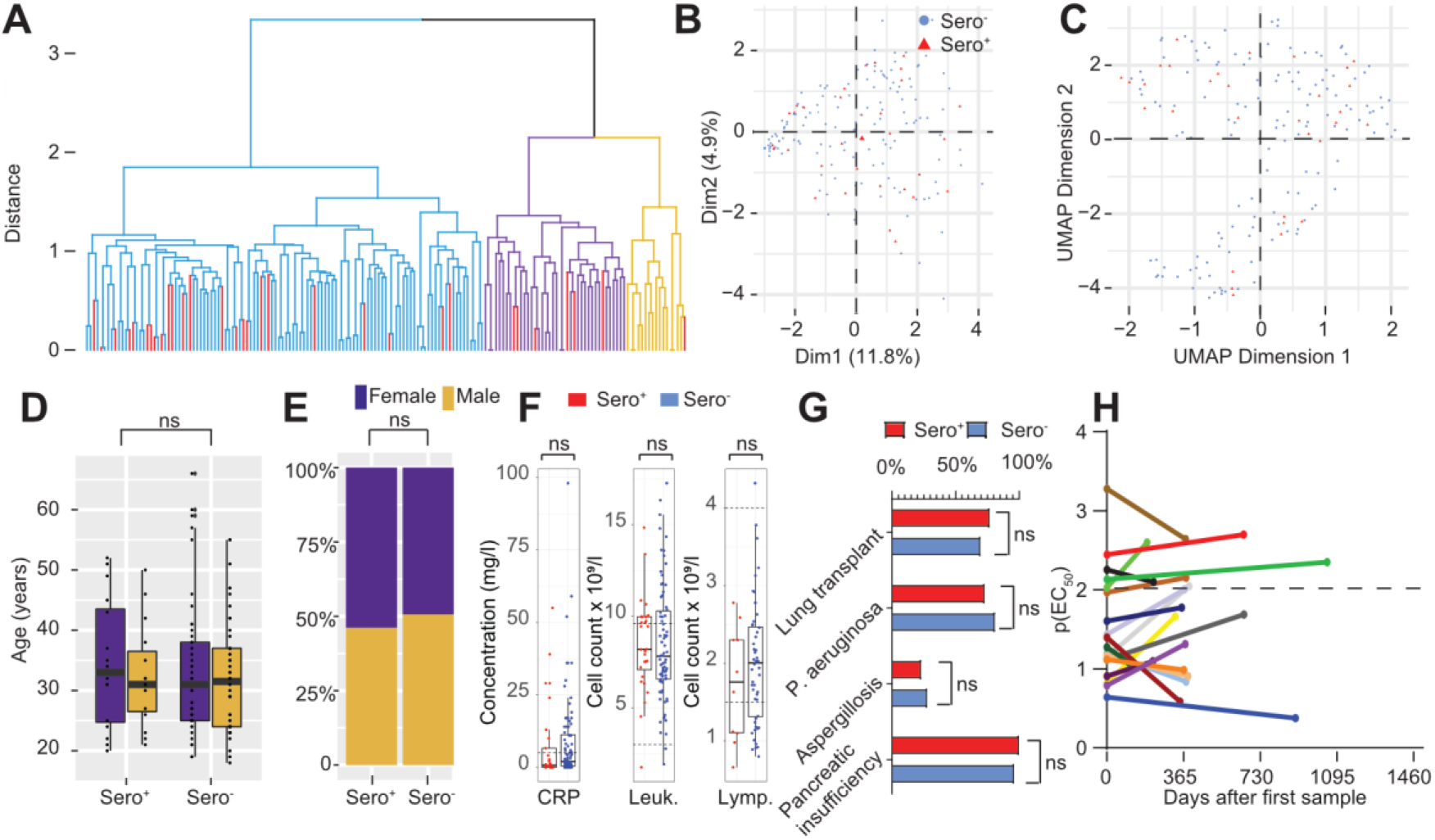
No clinical difference between seropositive and -negative pwCF. **A**. Hierarchical clustering analysis of ICD-10 codes and sex of seropositive and seronegative pwCF. Red bars: seropositives. **B-C**. PCA (B) and UMAP analysis (C) of ICD-10 codes and sex of seropositive and seronegative pwCF. **D-E**. No difference in age distribution (D, Mood’s median test) and sex (E, Χ^2^) of seropositive and seronegative pwCF statistics. **F**. No significant differences in CRP (mg/l), leukocyte and lymphocyte count (x10^9^/l) between seropositive and seronegative pwCF (Wilcoxon rank sum test with Benjamini-Hochberg correction). Dashed lines: reference range. **G**. No significant differences in clinical CF hallmarks between seropositive and seronegative pwCF (Fisher’s exact test). **H**. Temporal evolution of p(EC_50_) in 19 seropositive pwCF.

For 19 pwCF, we could measure two blood samples taken at different time points at least 104 days apart. The resulting p(EC_50_) values between these two samples were highly replicable (Pearson correlation coefficient R = 0.93, **Fig. S1D**), suggesting that the presence of such IgG antibodies is a stable phenomenon. We then plotted the temporal evolution of p(EC_50_) reactivity in these matched samples (**Fig. 3H**). Although the number of samples available was limited, the anti-Arah h 2 antibody seropositive individuals with CF stayed mostly seropositive, and the seronegatives stayed seronegative over the period investigated (up to 3 years). Hence, using the parameters at hand, we were unable to cluster seropositive and seronegative pwCF based on salient features of CF pathology.

### Antibody isotypes, subclasses, and affinity of antibodies against nAra-h2

Immune responses to food allergens in allergic patients are dominated by Th2-dependent priming of B cells into IgE-secreting plasma cells (Rogosch et al., 2010) which disrupts the tolerogenic local IgA-driven response (Berin, 2012; Tordesillas et al., 2017). However, patients with peanut allergy can develop both IgE and IgG antibodies against peanut allergens (El-Khouly et al., 2007; McKendry et al., 2021; Tay et al., 2007). We therefore characterized the antibody response to nAra-h2 in selected seropositive pwCF of whom we were able to obtain a large blood sample by means of patient recontacting. Peanut-allergic patients and healthy donors served as controls.

First, we conducted an IgG ELISA, with nAra-h2 coated to microplates, to re-evaluate the results obtained in the HTS. While pwCF displayed high p(EC_50_) values against nAra-h2, peanut-allergic patients were found to only have moderate IgG titers, and no reactivity was found in healthy controls (**Fig. 4A**). One patient with CF (CF5), whose previous sample showed a high titer against Ara-h2, did not show anti-Ara-h2 reactivity above threshold. Serum IgA against nAra-h2 was detectable in only two pwCF and in one patient with peanut allergy (**Fig. 4A**). To determine nAra-h2-specific serum IgE antibodies, we used the commercial ImmunoCAP technology, an automated sandwich fluorescence immunoassay, with whole peanut extract as antigen (**Fig. 4B**). The peanut-allergic patients, but none of the pwCF or patient controls, displayed high levels of IgE antibodies against peanut extract, with some of them exceeding 100 kU_A_/I and all falling into CAP class 3 or higher, indicating at least moderate and mostly high allergen sensitisation. We then investigated IgG subclasses using a panel of previously validated secondary antibodies (Emmenegger et al., 2022) in the same patients. Both IgG1 and IgG2, but not IgG3, showed minor contributions to the overall IgG response in pwCF, whereas the strength of the response was governed by the IgG4 subclass (**Fig. 4C**-**D**), which is remarkable as this phenotype is usually observed after allergic desensitization (M. Akdis & Akdis, 2014; Couroux et al., 2019).

**Fig. 4.**
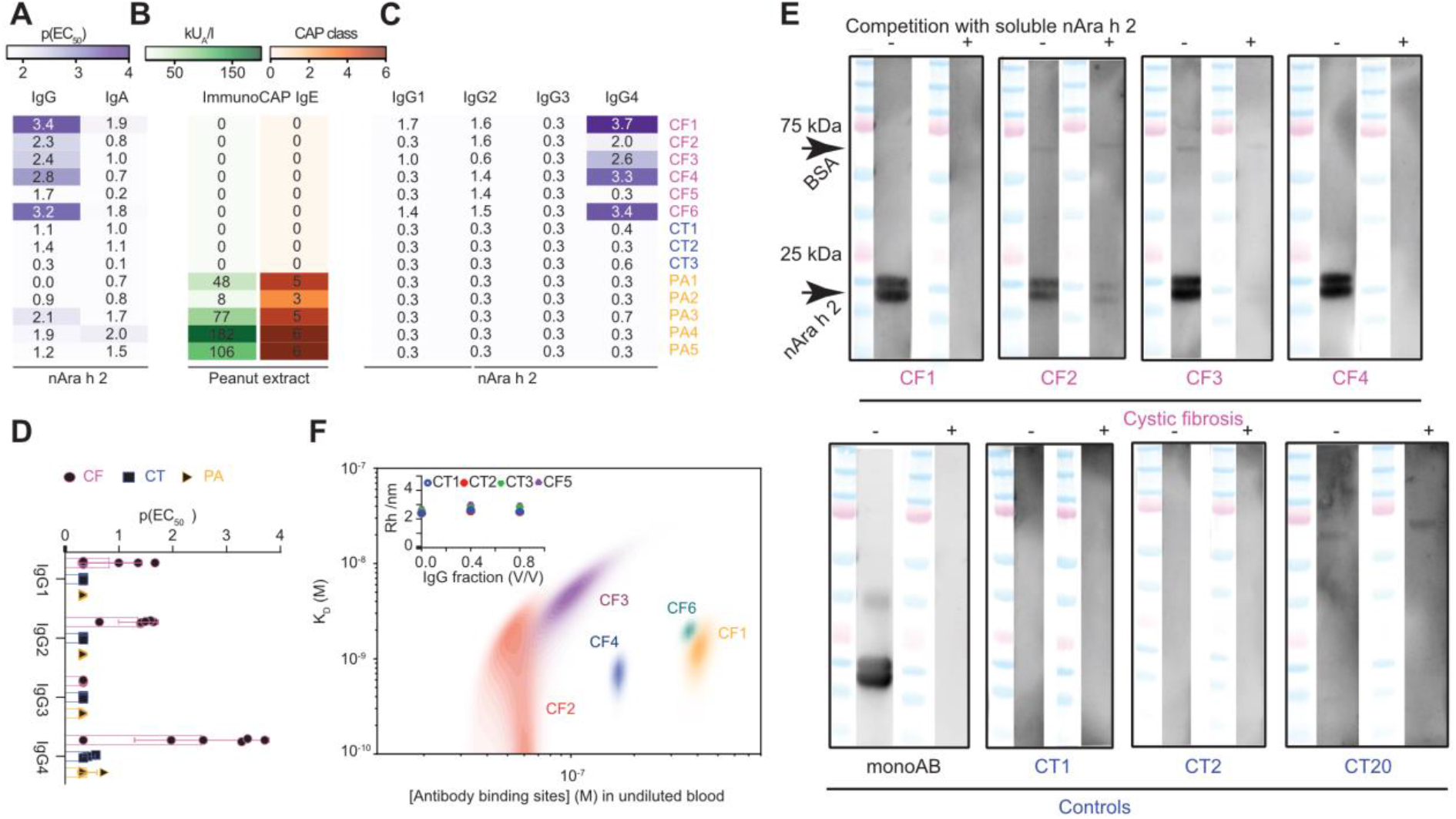
Analysis of the antibody response against Ara-h2 in PwCF and in controls. **A**. Analysis of the IgG and IgA response against Ara-h2 in six pwCF, three control patients (CT), and five peanut allergy (PA) patients. **B**. Analysis of the IgE response against peanut extract using ImmunoCAP. **C**. Analysis of IgG subclasses. **D**. Representation of data shown in (C) as boxplot with individual dots. **E**. Competitive Western Blot analysis. In each lane, an electrophoresis with nAra-h2 and BSA was conducted.

Next, we characterized samples by Western blotting. Strips containing a mix of Ara-h2 and BSA were incubated with plasma samples at a dilution of 1:100, optionally pre-incubated with soluble nAra-h2 for competition. PwCF, but no controls, showed the expected signal for nAra-h2 at 17-19 kDa in the non-preincubated condition. The signal almost entirely disappeared when the sample was pre-incubated with soluble nAra-h2, except for CF patient #2 (**Fig. 4E**), suggesting high-affinity interactions at low antibody concentrations.

To characterize the affinity of antibodies against nAra-h2, we used microfluidic diffusional sizing (MDS) (Emmenegger, De Cecco, et al., 2021; Emmenegger, Worth, et al., 2023; Schneider et al., 2022, 2023) on purified IgG fractions of plasma samples. The affinity and concentration profiles were determined for all pwCF except CF5 which did not display an increase in hydrodynamic radius even after concentrating IgG. For th3e samples for which a maximum likelihood value for the affinity was measurable, the measured affinity ranges were 0.75-3.93 nM, and the concentrations in undiluted serum were 0.02-0.1 μM (**Fig. 4F**). One sample (CF2) did not give a maximum likelihood value for the affinity, but the *K*_d_ was below 7.69 nM. We conclude that the anti-Ara-h2 IgG antibodies contained in the plasma of pwCF neutralize Ara-h2 with high affinity *in vitro*.

### pwCF develop IgG against certain food allergens but not pollen allergens

Next, we investigated whether pwCF harbour antibodies against multiple food allergens beyond Ara-h2. We selected 177 pwCF samples (median age 34 (IQR: 26-42) years, female:male ratio = 51:49) from our biobank and 76 samples from pediatric pwCF (median age 13 (IQR: 9-17) years, female:male ratio = 45:55) from a geographically distant locale. Thus, we introduced a validation cohort. For controls, we randomly selected 319 samples from hospital patients (median age 54 (IQR: 39-67) years, female:male ratio = 48:52), and from 358 healthy donors (median age 44 (IQR: 31-55) years, female:male ratio = 60:40) from the blood donation services of the Canton of Zurich (**Table 1**). These samples were tested for the presence of IgG antibodies against some of the common food allergens, here nAra-h1, nAra-h2, nAra-h3, nAra-h6, *Corylus avellana* (Cor-a) 9, *Glycine max* (Gly-m) 5, Gly-m 6, *Daucus carota* (Dau-c) 1, *Malus domestica* (Mal-d) 1, *Bos domesticus* (Bos-d) 11, *Betula verrucosa* (Bet-v) 1, and *Lolium perenne* (Lol-p) 1 using our HTS platform. As further controls, we used LAG3 extracellular domain containing a his-tag (Emmenegger, De Cecco, et al., 2021) and an uncoated condition to account for spurious binding to the plastic of the plate. The respective p(EC_50_) values were plotted as heatmap (**Fig. 5A**). The IgG reactivity of pwCF, but not of controls, against nAra-h2 was confirmed. Multiple additional epitopes displayed binding, including Ara h proteins, nGly-m5, nGly-m6, and nCor-a9. Using a cut-off p(EC_50_) value of 2, we classified individuals as seropositive or negative, and plotted the positive samples by condition (**Fig. 5B**). Among those showing consistently significant changes in adult and pediatric pwCF (**Fig. 5B**, **Supplemental File 06**), 4.5-10.5% of pwCF but only 0.6-1.4% of patient controls or healthy donors had antibodies against nAra-h1 (p: < 0.0365, Fisher’s exact test), 19.7-19.8% of pwCF but only 0-0.3% of controls had antibodies against nAra-h2 (p: < 0.0001, Fisher’s exact test), 26-31.6% of pwCF and only 1.3-2.8% of controls had antibodies against nAra-h3 (p: <0.0001, Fisher’s exact test), 21.1-23.2% of pwCF and 0% of controls had antibodies against nAra-h6 (p: <0.0001, Fisher’s exact test), 11.8-16.9% of pwCF and 0.6-1.1% of controls had antibodies against nGly-m6 (p: <0.0001, Fisher’s exact test). Importantly, IgG reactivity against the two pollen allergens was almost entirely absent in adult or pediatric pwCF (p: >0.999 for nBet-v1) or lower (adult) or equal (pediatric) to controls (for nLol-p1), indicating that allergens are not generically targeted by IgG antibodies in CF.

**Fig. 5.**
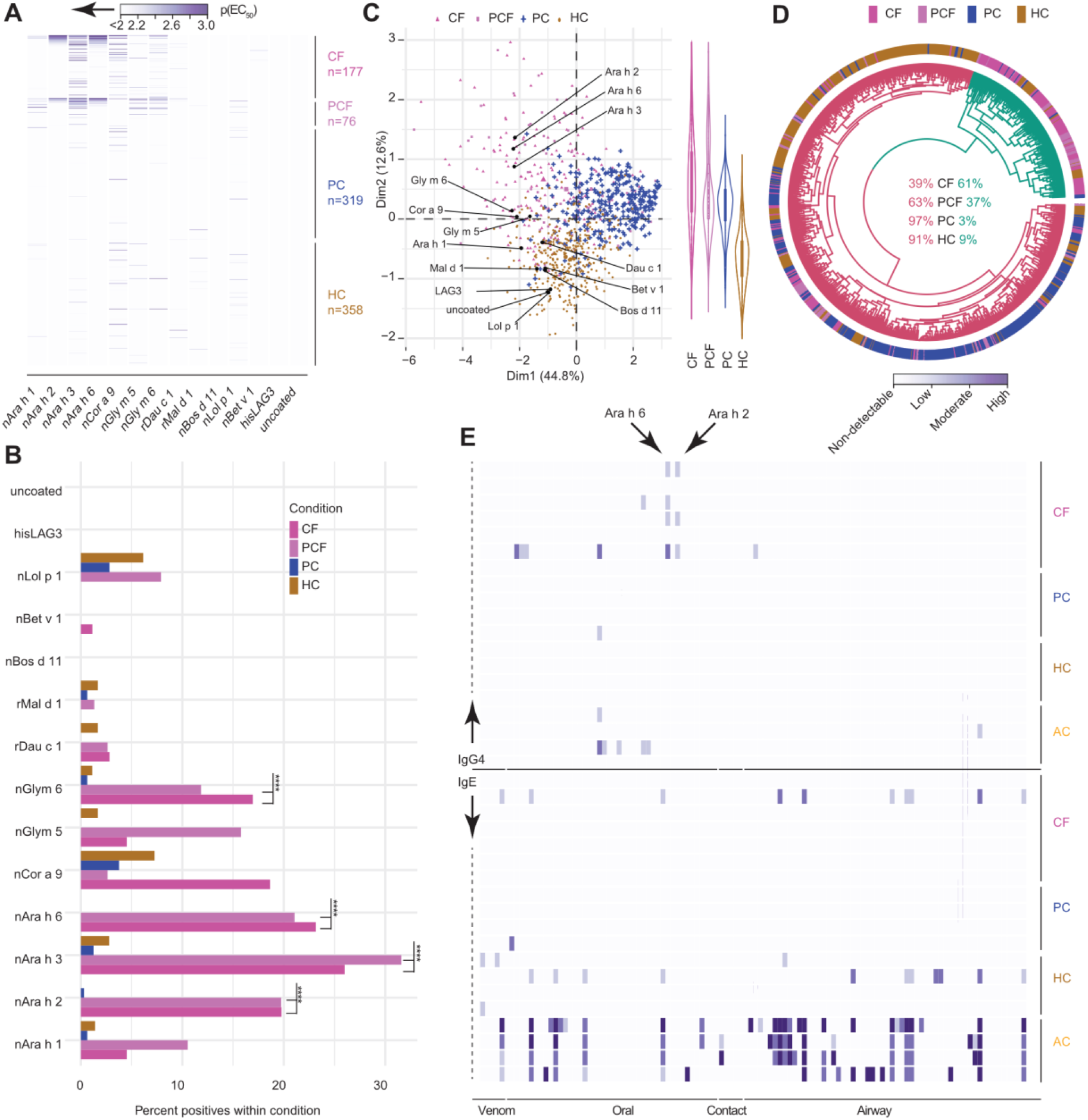
pwCF have IgG against food allergens but not pollen allergens. **A-B**. Heatmap (A) and percent positives (B) of IgG responses against select allergens and control proteins in adult (CF) and paediatric (PCF) pwCF, non-CF patients (PC) and healthy blood donors Fisher’s exact test. Detailed statistics in **Supplemental File 06**. **C**. Two-dimensional representation of data from (A) after PCA. Right side: violin plot displaying the distributional differences of the second PCA dimension according to patient groups. **D**. Circularized unsupervised hierarchical clustering suggests that the dataset (from A) can be classified into two main groups, shown in green (predominantly adult or paediatric pwCF) and red (mostly non-CF controls). **E**. IgG4 (top) and IgE (bottom) reactivity in large allergen panel in subset of pwCF and controls, assessed using the ISAC technology.

The IgG profiles of pediatric and adult pwCF were similar (**Fig. 5A**-**B**) with slight differences for nCor-a9, nGly-m5, and nLol-p1. A PCA-based cluster analysis performed on the p(EC_50_) values on all patients and antigens indicated that Ara-h2, Ara-h3, and Ara-h6 built a cluster mainly driven by adult (purple triangles) and pediatric (lila squares) pwCF (**Fig. 5C**). The patient controls (blue crosses) and the healthy donors (brown dots) formed two overlapping yet partially distinct clouds. We performed circularised hierarchical clustering on the same data set and colored the two main branches of the dendrogram (red and green, respectively), aiming to find whether pwCF and controls fall into either of these two main branches (**Fig. 5D**). The distribution of pwCF was shifted towards cluster 1 (green, 61% adults and 37% paediatric pwCF) much more than the distribution of the controls (3% patient controls and 9% healthy controls), corroborating the PCA-based visualization.

We then investigated the allergen profiles with the ImmunoCAP ISAC technology, incorporating an array of 112 allergens from 48 sources, including food (oral), pollen (airway), venom, and contact allergens (Jakob et al., 2015; Matricardi et al., 2016). IgG4 and IgE antibodies were measured in a subset of pwCF and controls. Using a standardized cutoff of 0.3 international standard units (ISU), we observed that pwCF carry IgG4, but not IgE, antibodies against Ara-h2 and Ara-h6 (**Fig. 5E**), as shown with ELISA (**Fig. 5A-B**). In the same patients, there was no reactivity with allergens entering through the airways, an observation we substantiated with raw data where no cutoff has been applied (**Fig. S2** and **Supplemental File 07**). This is opposed to allergic patients displaying IgE reactivity against a multitude of airway, contact, food, or venom allergens. While the cutoff-based representation draws a clear picture due to its stringency (**Fig. 5E**), the raw value map indicates that more food allergens may be targeted by IgG4 in CF (**Fig. S2**), confirming our ELISA data.

We conclude that a subset of pwCF develops selective immunity to food proteins, in particular peanut allergens Ara-h2 and Ara-h6, at much higher levels than controls. Importantly, our observation that healthy controls harbor serum IgG against multiple food proteins expands the finding that certain food antigens elicited systemic IgG in up to 50% of individuals (Leviatan et al., 2022).

### Lack of evidence for cross-reactive antibodies and absence of broad reactivity against endogenous antigens in pwCF

We addressed the possibility that reactivity is governed by cross-reactive antibodies primarily matured to neutralize infectious pathogens. A protein blast using BLOSUM-62 matrix suggested various homologies of Ara-h2 with seed storage proteins of related plant species (e.g. with Ara d 2 of Arachis duranensis, Ara h 6 of Arachis hypogaea, with related conglutinins of Lupinus albus and angustifolicus, or with albumin of Glycine max or Punica granatum, see **Supplemental File 08**), and previous structural investigations have not revealed similarities beyond the kingdom of plants (Mueller et al., 2011).

We further investigated the immune profile in pwCF by Rapid Extracellular Antigen Profiling (Wang, Dai, et al., 2021). Using samples from adult and pediatric pwCF and of a range of controls, we subjected yeast cells transfected with an array of 6,184 plasmids encoding transmembrane and extracellular proteins (**Supplemental File 09**) to the purified IgG fraction of the above patients and calculated the fold-change in detection, suggestive for antibody binding. We then selected proteins with a log2-fold increase of ≥ 2 in at least one sample (n = 293, **Fig. 6A**) and excluded proteins upregulated (presumably nonspecifically) in all conditions (n = 53, see **Supplemental File 09**). No striking patterns among the different patient conditions emerged and no significant differences in the number of autoreactive antibodies in pwCF and controls was observed (one-way ANOVA, **Fig. S3**). A PCA analysis of the above dataset showed no clusters in adult and paediatric pwCF (green) as well as controls (yellow) (**Fig. 6B**), although some adult and paediatric pwCF showed a tendency towards higher PC1 and negative PC2 values. A separate feature cluster was formed by a group of alpha interferons (e.g. IFNA8, IFNA4) that appeared as positive in two control patients but not in pwCF (**Fig. 6A**-**B**), with IFNAs being important contributors to the second PC (blue dots) as measured by the squared cosine (cos2) function (Abdi & Williams, 2010). Conversely, the dataset is suggestive of an enrichment of autoantibodies targeting the type III interferons IFNL2/IL28A and IL28B, important contributors to the first PC, in some pwCF. To analyse these patterns using an alternative approach, we mapped all data (n = 6,184), including the nonspecifically upregulated ones, in a group-wise fashion, displaying the difference of the log2-fold change of pwCF and controls as a function of the average log2-fold-change in all individuals. All but five proteins are within 2 log-fold changes (from -1 to +1), suggesting no aberrant autoreactivity in pwCF compared to controls (**Fig. 6C**). IL28A, IL28B, and IL29 appeared to be enriched in pwCF, indicating that autoreactivity against type III interferons might be increased. This data suggests a normal autoimmune profile in CF, judged by the protein antigens that were employed with this technology, as opposed to previous investigations on systemic lupus erythematosus (Wang, Dai, et al., 2021) or COVID-19 (Wang, Mao, et al., 2021). On the other hand, CF has been associated with elevated IgA autoantibodies against double-stranded DNA (Yadav et al., 2020) and with anti-PAD4 autoantibodies (Linnemann et al., 2022).

**Fig. 6.**
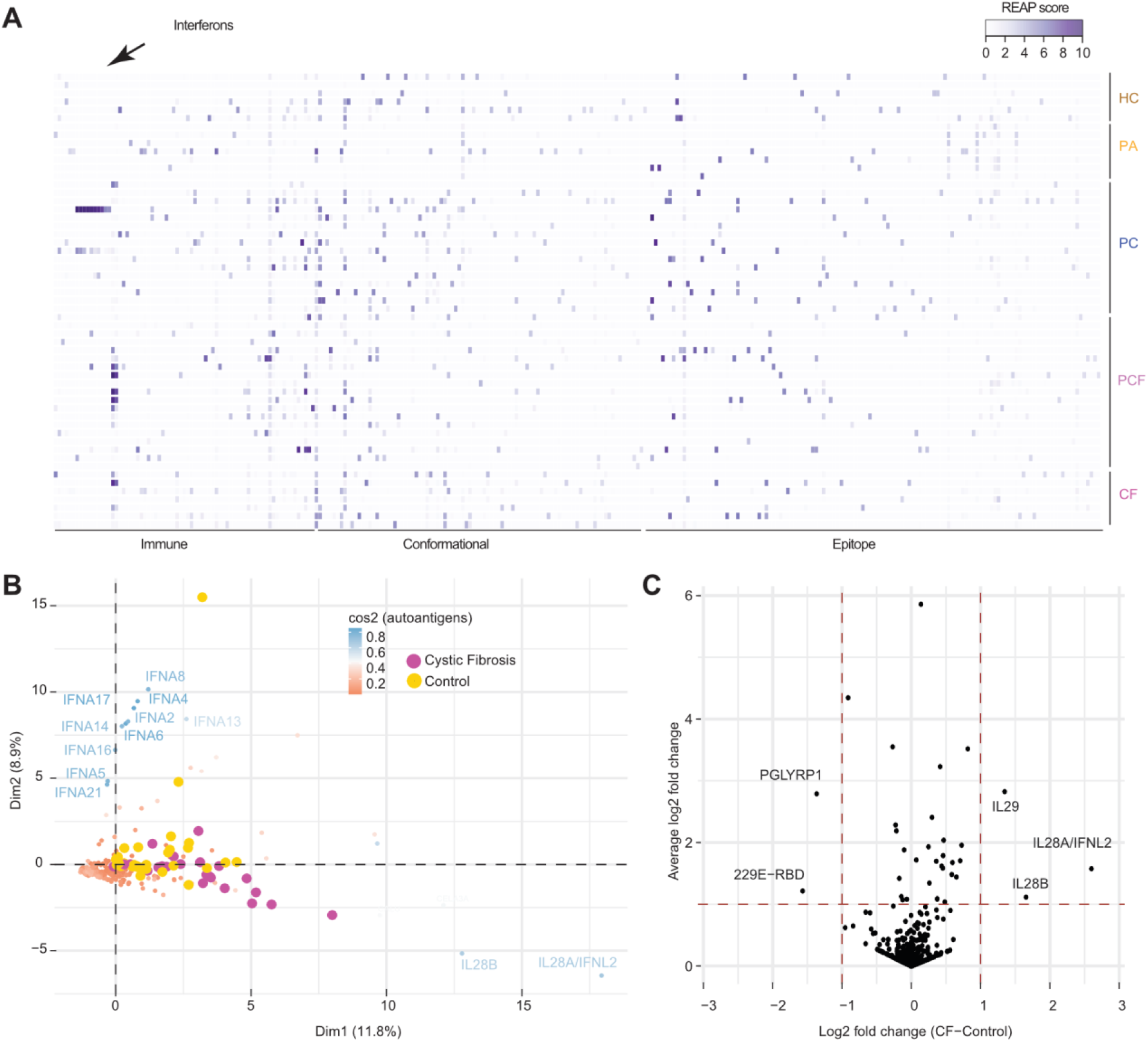
No evidence for cross-reactive antibodies and absence of aberrant autoimmunity profile in pwCF. **A**. REAP analysis. Shown are proteins with a log2-fold increase of ≥ 2 in at least one sample. **B**. PCA representation of the data shown in (B). Data is grouped for pwCF and non-CF controls. **C**. Average log2-fold-change per group/log2-fold-change by all samples, grouped for pwCF and non-CF controls. B and C. Autoantibodies against type III interferons (IL28A/B and IL29) show a slightly altered profile in pwCF versus controls.

### No marked changes in B cell receptor repertoires of pwCF, peanut allergic, and healthy donors

To investigate the entire antibody repertoire in an unbiased manner, we performed adaptive immune receptor repertoire (AIRR) sequencing (Miho et al., 2018; Rubelt et al., 2017) in anti-Ara-h2 seropositive pwCF (n = 6), patients with peanut allergy (n = 3), and in healthy donors (n = 3). Sequencing of the variable heavy (VH) and variable light (VL) BCR repertoire from each subject produced >10^6^ raw reads for most samples (**Fig. S4A**). After quality control and processing, a high-fidelity data set consisting of unique error-corrected IgM, IgD, IgG, IgA, and IgE sequences and unique error-corrected IgL and IgK sequences was generated (**Fig. S4B**-**C**). The pwCF showed the highest percentages of IgA sequences among the total VH repertoire (44.5%, with IgG contribution of 28.1%), whereas the VH repertoire of the peanut allergy group was composed of predominantly IgG (38.6%), IgA (29.1%) and to a lesser extent, IgD (5.4%). The peanut allergy group isotype repertoire contained some IgE sequences (0.23% on average), while it was almost completely absent in pwCF (0.023% on average). IgM was the largest component of the healthy VH repertoire (41.0%) while IgG (29.3%) and IgA (19.8%) followed. None of the distributions showed significant differences between patient groups (pairwise t-test). In line with comparisons among isotypes, the heavy and light chain distributions were not significantly different among the three patient groups (**Fig. S4C**, pairwise t-test). As expected, VH3 family usage was more pronounced for all the Ig isotypes and patient groups, followed by VH4, VH1, with hardly any contribution by VH2 and VH5. Statistical comparisons between patient groups did not reveal any significant difference (**Fig. S4D**).

We analyzed the abundance of antibody variable domains to test whether the different isotype compartments in CF or peanut allergy resulted from clonal expansion. Sequences that differ only by isotype were clustered into groups consisting of expanded clones, where the size of each clone is determined by the number of sequence members (Vander Heiden et al., 2017). The largest clone size in the CF group reached 6.18% (95% CI: 1.61-10.7%) for IGHG (**Fig. 7A**), suggestive of an active immune response (Laserson et al., 2014). Clone size in the IGHA, IGHD, and IGHM compartments of all patient groups did not exceed 5% of the B cell repertoire. IGHE clones were expanded in peanut-allergic patients (but not in any other group), exceeding 20% of the total BCR repertoire.

**Fig. 7.**
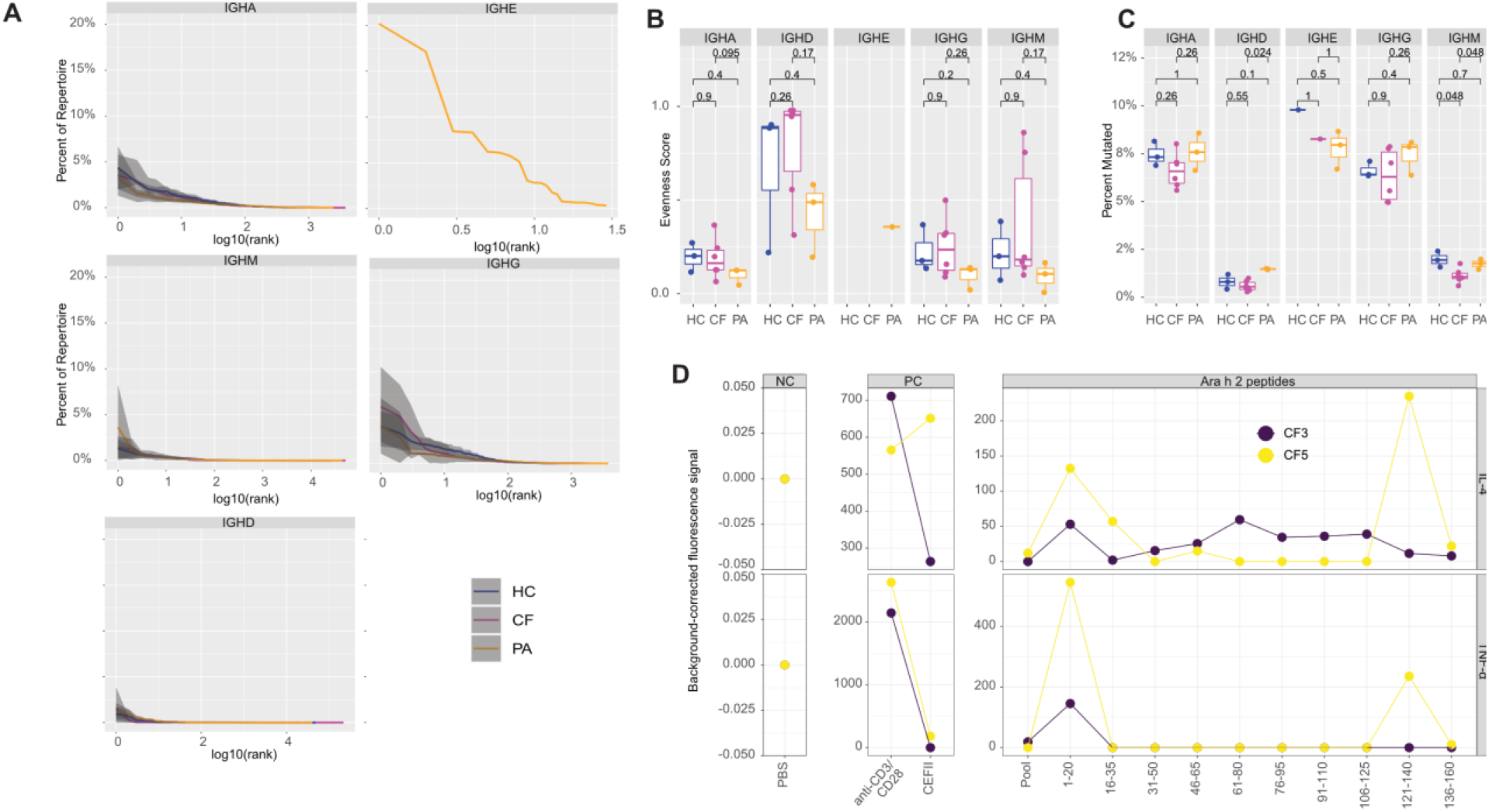
B cell receptor repertoire sequencing and T cell activation assay. **A**. Abundance in antibody variable domains suggests no significant differences among the isotype compartments in pwCF, peanut allergy or controls. **B**. No statistical differences in the evenness score were identified among the groups, suggesting that clonal expansion follows the same pattern in pwCF compared to other patient groups. Wilcoxon rank sum test. **C**. The mutational frequency was high for the class-switched IGHA and IGHG but comparatively low for the non-class-switched IGHM and IGHD, as expected. No statistical differences among the groups were identified. Wilcoxon rank sum test. **D**. T cells of two seropositive pwCF activated with pooled Ara-h2 peptides or with individual Ara-h2 peptides release IL-4 and TNF-α. PBS was used as negative control and anti-CD3/CD8 antibodies and CEFII as positive controls.

By calculating the variance of the abundance of the clones we also estimated the evenness of the CF, peanut allergy, and healthy donor groups. Evenness serves as a measure of clonal dominance; samples with low evenness would have a less uniform distribution and would therefore be characterized by fewer but highly expressed clonotypes, whereas samples with similarly abundant clones would have higher evenness. The evenness was inferred from the Hill diversity index (Hill, 1973) calculated over a range of diversity orders (q) ranging from 0 to 4, in equally spaced increments, for each subset **(Fig. S5A**). The evenness score tended to be slightly higher in CF than in the two other groups for IGHG, indicating that expansion was more diverse and not bound to few clones (**Fig. 7B**). However, none of the group-wise comparisons reached statistical significance, suggesting that there were no profound differences in the BCR repertoire between the three groups.

To determine whether somatic hypermutation (SHM) is perturbed in CF, we analyzed somatic mutation patterns of IGHV gene sequences. SHM is known to be biased toward DNA motifs prone to mutations (hotspots). Mutation frequency for each sequence was calculated as the number of base changes from germline in the V region. Mutational frequency displayed isotype-specific differences. As expected, the mutational frequency was high for the class-switched IGHA and IGHG but comparatively low for the nonclass-switched IGHM and IGHD. IGHE appeared at medium mutational frequency; however, the number of sequences were low (**Fig. 7C** and **Fig. S5B**). Group-wise comparisons among the IGH isotypes did not reveal any significant difference, suggesting that pwCF do not have a broadly altered rate of SHM.

In summary, AIRR sequencing revealed that the antibody repertoire in pwCF does not substantially deviate from those observed in healthy controls or patients with peanut allergies with regard to isotype composition, clonal expansion, clonal dominance, and SHM. This finding further highlights the specificity of the IgG response against peanut allergens identified in pwCF in the absence of global alterations of the BCR repertoire. In the light of the many chronic conditions connected to CF, including the pwCF investigated in this study, e.g. chronic infections and immune suppression, the scarcity of difference in the BCR repertoire is surprising.

### Absence of CDR3 convergence between sequences obtained via BCR repertoire sequencing of pwCF and publicly available clones known to bind Ara-h2

Since peanut allergy and pwCF both have antibodies targeting peanut allergens, we hypothesized that there might be common clones with identical or highly similar CDR3 regions that diverged from the germline sequence. Common clonal groups have been found among unrelated peanut-allergic patients that have been treated with oral immunotherapy (OIT) (Kiyotani et al., 2018; Patil et al., 2015). Additionally, common clones of the IgE isotype were also detected in untreated peanut-allergic patients with identical gene rearrangements within VH and VL chains (Croote et al., 2018). Thus, we aligned the heavy and light chain CDR3 regions of our cohorts to publicly available sequences known to recognize Ara-h2 retrieved from several studies (Aguzzi et al., 2020; Croote et al., 2018; Hoh et al., 2016). We identified a small number of convergent antibody responses across all disease and control groups that did not exceed 0.2% of the heavy chain CDR3 sequences (**Fig. S6**). As expected, light chain convergence was higher reaching 15% of the CDR3 kappa chain and no more than 10% of the CDR3 lambda chain across all groups since light chain heterogeneity is low in the general population (Collins & Watson, 2018). There was no evidence for convergent antibody responses in the BCR repertoire in pwCF and peanut allergy.

### T cells are activated with Ara-h2 peptides

Several studies did not show a clear association between peanut allergy and HLA (Dreskin et al., 2010; Shreffler et al., 2006) whereas others showed a higher frequency of DR8 (Boehncke et al., 1998; Howell et al., 1998). In addition, the HLA-DQ gene region, especially DQ6, was linked to peanut allergy in two comprehensive studies (Hong et al., 2015; Madore et al., 2013). HLA studies performed on pwCF revealed increased frequencies of DR4 and DR7 alleles compared to controls (Aron et al., 2012), however, findings were not reproduced in a large study (Adriaanse et al., 2014). Here, we characterized the human leukocyte antigen (HLA) class II locus of pwCF with IgG reactivity to peanut allergens. This approach was governed by the assumption that the high-affinity B cell responses observed in this collective (**Fig. 4**) require T cell help. The identification of T cell epitopes may provide further explanation for the mechanism by which some pwCF develop an adaptive immune response to certain food allergens.

We genotyped HLA-class II alleles DRA*, DRB1*, DRB3*, DRB4*, DRB5*, DQA1* and DQB1*, DPA1*, and DPB1* in pwCF 1-6 (for summary of results, see **Supplemental File 10**). Certain HLA-DRB1* and -DRB3* alleles (DRB1*03:01:01G in 50%; DRB1*11:01.01G in 50%; DRB3*01:01:02G in 67% and DRB3*02:02:01G) and even more certain DQA* (DQA1*01:02:01G in 67% and DQA1*05:01:01G in 100%) and DQB1* (DQB1*02:01:01G in 50%, DQB1*03:01:01G in 50% and DQB1*06:02:01G in 50%) and DPA1* (DPA1*01:03:01G in 100%; 4/6 patients homozygote for this allele) and DPB1* alleles (DPB1*04:02:01G in 50%) were found at much higher frequencies than in the normal European population. Particularly the frequency of HLA-DQA1*05:01 was elevated 2-6-fold compared to control populations of similar ethnicity (Gonzalez-Galarza et al., 2020)). Moreover, the comparison of the amino acid sequences of HLA- DQA1*05:01:01 with the DQA1*01:01:01 allele and other DQA1* alleles of the six individuals revealed changes in size, charge and hydrophobicity at multiple positions that participate in forming the HLA-DQ binding groove, but also outside (see **Supplemental File 11**), and HLA-DQA1*05:01:01 differs most from the other alleles (Barker et al., 2023).

Finally, we asked if T-cell reactivity against overlapping peptides of the main peanut allergen Ara-h2, against which the antibodies are directed, provides further insight. We used Fluorospot testing of whole PBMCs with cytokines IL-4 and TNF-α as readout and included PBS as a negative and anti-CD3/CD28 antibodies as well CEFII, a mixture of peptides derived from cytomegalovirus, Epstein Barr Virus and influenza A virus, as positive controls. Two seropositive pwCF, for whom sufficient PBMCs were available, responded to multiple Ara-h2 peptides, with respective peaks at peptides corresponding to amino acids 1- 20, 61-80, and 121-140 (**Fig. 7D**). When performing in silico peptide binding predictions of Ara-h2 to the HLA-DQA/B combinations DQA1*05:02/DQB1*02:01:01 and DQA1*05:02/DQB1*03:01:01 (NetMHCII-pan-4.0; see (Jensen et al., 2018)), no strongly or weakly binding peptide was identified in the Ara-h2 sequences considered important in the cellular assay (see **Supplemental File 12**), and therefore, strong HLADQ binding of this region is unlikely.

### CFTR heterozygosity is not associated with peanut allergy or food allergy in general

Mutations in the CFTR affect not only epithelial cells but also macrophages and lymphocytes (Bruscia & Bonfield, 2016, 2022; Hartl et al., 2012) of heterozygous CFTR mutation carriers (Miller et al., 2020; Polgreen et al., 2018). We wondered if CFTR heterozygous carrier states may be associated with altered antibody responses against peanut and other food allergens. We examined an anonymized collective of confirmed peanut-allergic patients (n=90) for the presence of mutations in the *CFTR* gene and compared it with the expected frequency in the non-Finnish European population. We restricted the analysis to variants with available information contained in gnomad3.1.1 and additionally displayed data from the CFTR2 database (**Fig. 8A**). The cumulative allele frequency, which correlates with heterozygosity (**Fig. 8B**), was 4.0% for peanut-allergic patients, almost identical with the population (4.1%, according to gnomad3.1.1) and similar to the non-Finnish Europeans (3.3%), while the allele frequency of pwCF, based on data from CFTR2, was lower (1.2%). We then displayed the allele frequencies for all variants (**Fig. 8C**). We observed two significant alterations for variants c.743+40A>G (p=0.005, t-test) and for c.1585-1G>A (p<0.0001, t-test). The c.1521_1523del (p.Phe508del) mutation, which is the most common cause of CF (69%), has an allele frequency of only 1.4% (non-Finnish European population) and 1.1% (peanut-allergic patients).

**Fig. 8.**
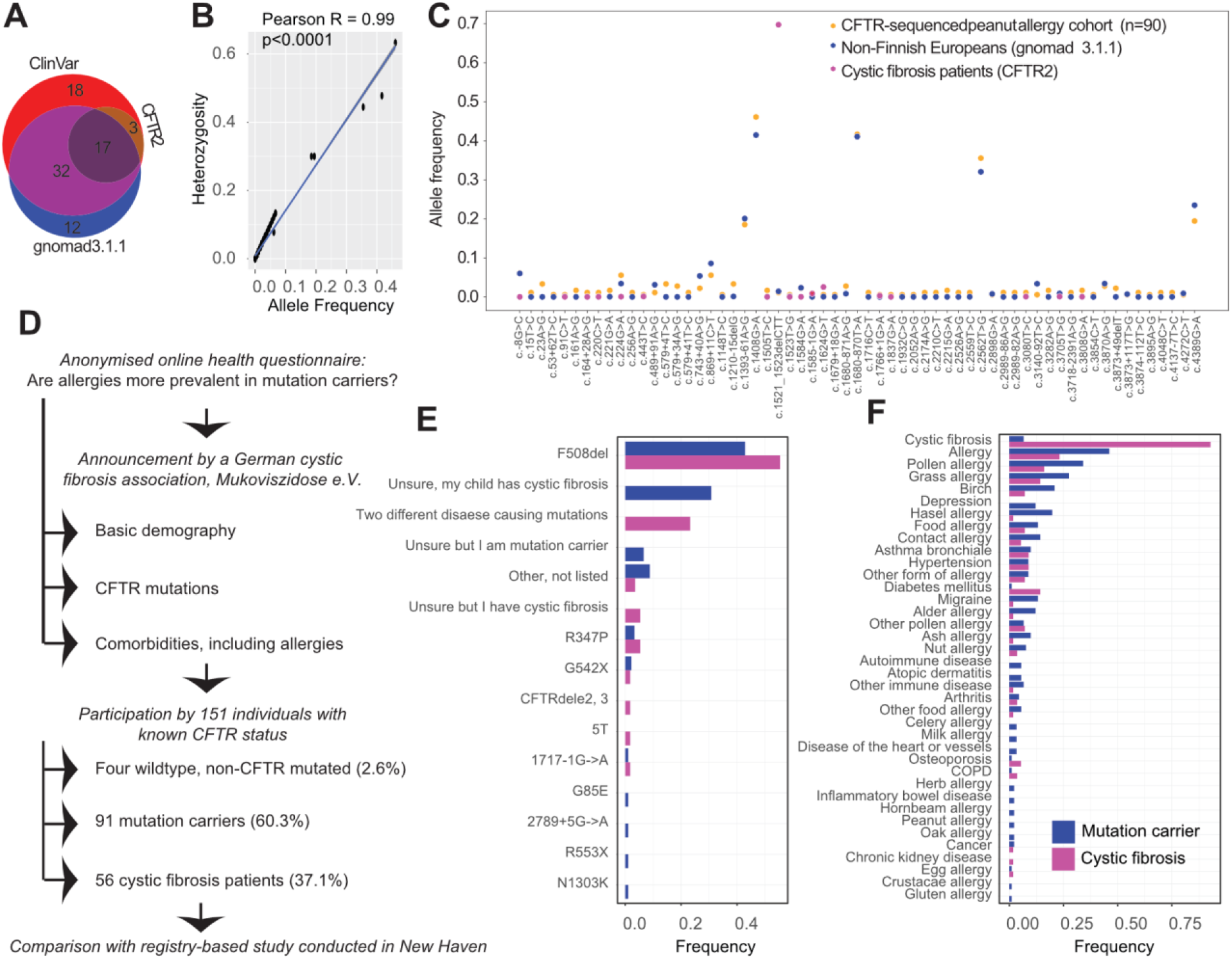
CFTR sequencing in patients with peanut allergy and health survey. **A**. Repositories used to analyze and compare the data. **B**. Cumulative allele frequency correlates with heterozygosity. **C**. Allele frequencies for all CFTR variants in CFTR-sequenced PA cohort (n=90), and in controls. **D**. Health surveys assessing the frequency of food allergies in pwCF compared with heterozygous controls in Europa and in North America. **E-F**. Frequency of mutations (E) abd health conditions (F) as indicated by study participants, for mutation carriers and pwCF (logistic regression analysis).

We then invited individuals with known CFTR status (wildtype, mutation carrier, or pwCF) to participate to an anonymized survey on medical symptoms including allergies (**Fig. 8D**). The survey was distributed online by the German CF association “<u>Mukoviszidose e.V.</u>” and was broadly accessible without restriction. It queried basic demographic information and a panel of comorbidities including allergy (**Supplemental File 13**). 153 individuals participated between July 2022 and January 2023, of which two did not include any information and were excluded. Among the 151 individuals, four (2.6%) indicated to not carry any CFTR mutation. 91 individuals (60.3%) were heterozygous mutation carriers, and 56 individuals (37.1%) reported to have CF. The median age was 36 (IQR:16.5-43) years, with 62.3% of participants being females (**Table 3**). The majority of carriers (43%) and pwCF (55%) reported a deletion of Phe^508^; some (31%) did not know the details of their sequence while being parent of a child with CF, and 23% of pwCF were trans-heterozygous (**Fig. 8E**).

**Table 3.** Population characteristics for online survey investigating differences in the prevalence of allergy between CFTR mutation carriers and pwCF. The questionnaire was provided in German language and announced by a German CF association.

|  | *Wildtype* | *Mutation carrier* | *Cystic fibrosis* | *Overall* |
| --- | --- | --- | --- | --- |
| *Individuals, number* | *4* | *91* | *56* | *151* |
| *Median age (IQR), years* | *24.5 (14.0-33.5)* | *40.0 (34.0-45.0)* | *13.5 (6.0-31.0)* | *36.0 (16.5-43.0)* |
| *Sex, female %* | *75* | *63.7* | *58.9* | *62.3* |
| *Sex, male %* | *25* | *36.3* | *41.1* | *37.7* |
| *Food allergy %* | *0* | *13.2* | *7.1* | *10.6* |

As the number of non-mutated individuals was too low for a statistical comparison, we directly compared the prevalence of medical conditions between mutation carriers and pwCF using multivariable logistic regression adjusted for age and sex. There was no significant difference regarding the prevalence of allergy in general (46.2% mutation carriers and 23.2% pwCF self-reported an allergy), or of subgroups thereof (the most frequent allergy type was pollen allergy, and 13.2% mutation carriers and 7.1% pwCF indicated they have some food allergy), between these groups (**Fig. 8F**), consolidating the result from the sequencing-based approach. The only two conditions significantly different between the two groups were CF (odds ratio: 132.6 (CI95%: 38.1-607.0), p <0.0001) and diabetes mellitus (odds ratio: 32.6 (CI95%: 5.1-645.0), p=0.0018). While this survey was conducted primarily in Germany, a registry-based investigation conducted at the Yale New Haven Hospital center in New Haven, USA, resulted in the same conclusions. In 115 individuals (median age 5.0 (IQR: 2.0-15.0), 48.7% of female sex) whereof 56 individuals were CFTR mutation carriers, and 59 individuals were pwCF, the prevalence of food allergy was non-significantly different (p=0.345, multivariable logistic regression adjusted for age and sex), with 7.1% mutation carriers and 8.5% of pwCF having a clinically diagnosed food allergy (see **Table 4**).

**Table 4.** Population characteristics for registry-based survey investigating differences in the prevalence of allergy between CFTR mutation carriers and pwCF. The survey was conducted in Yale, New Haven, United States of America.

|  | *Mutation carrier* | *Cystic fibrosis* | *Overall* |
| --- | --- | --- | --- |
| *Individuals, number* | *56* | *59* | *115* |
| *Median age (IQR), years* | *3.0 (1.8-4.0)* | *13.0 (8.0-18.0)* | *5.0 (2.0-15.0)* |
| *Sex, female %* | *44.6* | *52.5* | *48.7* |
| *Sex, male %* | *55.4* | *47.5* | *51.3* |
| *Food allergy %* | *7.1* | *8.5* | *7.8* |

Given the sum of evidence collected, we refute our initial hypothesis that peanut allergy or other food allergies are associated with heterozygosity in CFTR.

### Peanut sensitization in Cftr^-/-^ mouse model

We next assessed whether our finding could be recapitulated in a well-characterized murine model of CF (Snouwaert et al., 1992) which includes intestinal pathology. We speculated that intestinal dysfunction in CF (Collaco et al., 2010; Ooi & Durie, 2016), which includes aberrant mucus release (Garcia et al., 2009; J. Liu et al., 2015) and goblet cell hyperplasia with mucus accumulation in the crypt lumen and along the villous surfaces (Gorrieri et al., 2016; J. Liu et al., 2015; Walker et al., 2022), and increased gut permeability (De Lisle, 2014) combined with a state of chronic infections, may alter the immune reactions triggered by peanut allergens and prevent the development of allergy. Oral anaphylaxis to peanut has previously been shown to be associated with enhanced gut permeability (Gertie et al., 2022). *Cftr^-/-^* and wildtype (WT) mice (both C57BL/6) were orally immunized with ground peanut (5 mg/immunisation, with or without cholera toxin as adjuvant) for six weeks, allowed to recover for ten days, and subjected to an intraperitoneal challenge with 100 µg ground peanut (**Fig. 9A** for overview).

**Fig. 9.**
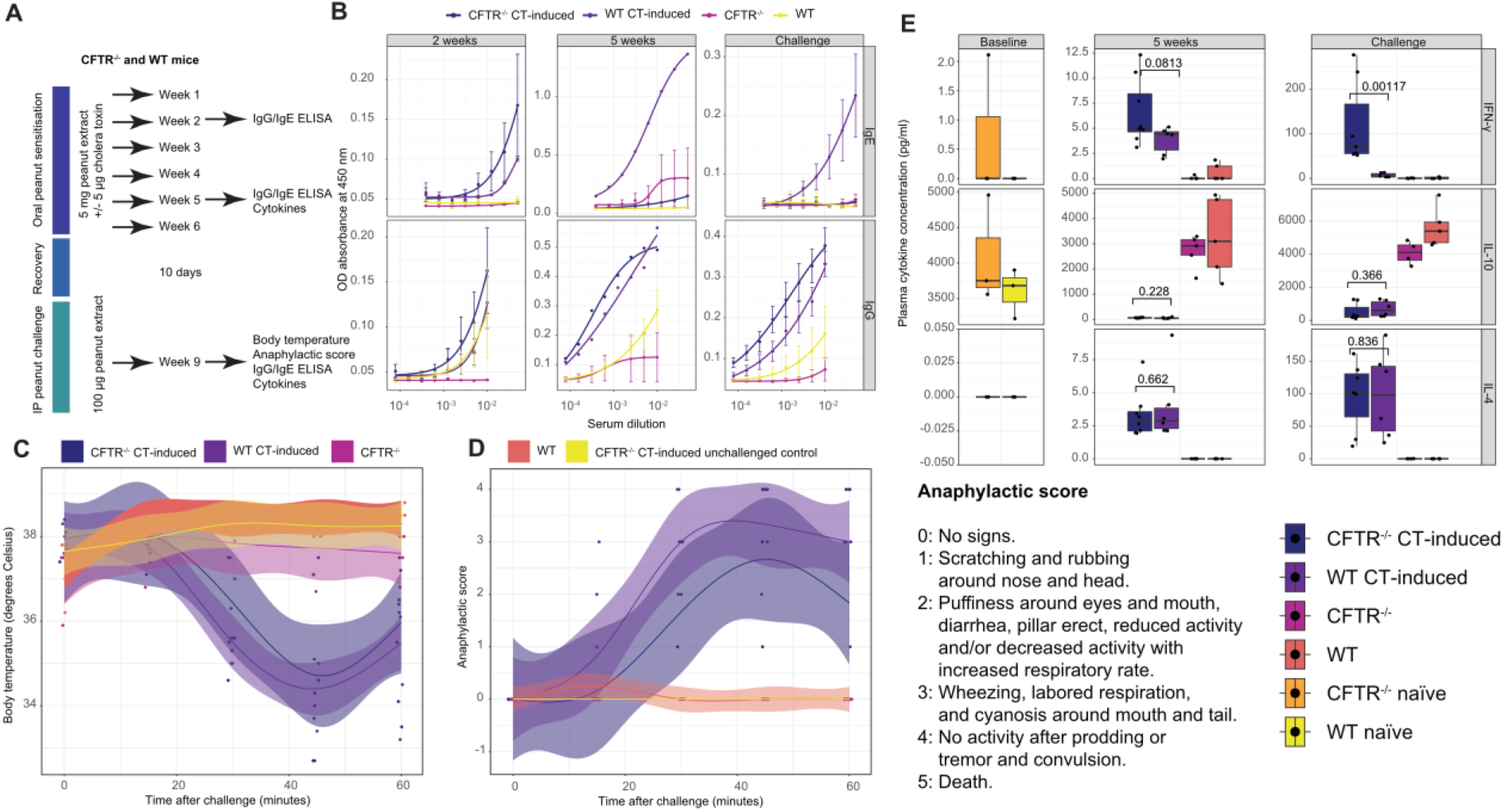
Peanut sensitisation in Cftr^-/-^ mouse model. **A**. Overview of peanut-allergic sensitisation experiments in *Cftr^-/-^* and wildtype mice. **B**. IgE and IgG ELISA in serum of *Cftr^-/-^* cholera toxin (CT)-induced mice of WT CT-inducted mice, *Cftr^-/-^* mice and WT mice at 2-5 weeks of sensitisation and post-challenge. IgE reactivity pattern is visible at 5 weeks and post-challenge. **C**. Change in body temperature measured in animals post intraperitoneal peanut challenge. **D**. Anaphylactic scores assessed in animals post intraperitoneal peanut challenge. Average shown with 95% confidence intervals. **E**. Cytokine profiles assessed at various timepoints in different mouse groups. IFN-γ levels were significantly different in *Cftr^-/-^* CT-inducted compared with WT CT-inducted mice post-challenge and the trend was already visible at five weeks. Wilcoxon rank sum test.

At week 2, modest IgG and IgE responses against crude peanut extract became visible in *Cftr^-/-^* mice and WT control mice (**Fig. 9B**). Antibody levels in cholera toxin-treated *Cftr^-/-^*mice were higher than in WT mice, likely due to an increased antigenic permeability of the intestinal barrier in connection with goblet cell hyperplasia. At week 5 and post challenge, we observed a marked IgE response in WT but not in *Cftr^-/-^*mice (p = 0.0019 and 0.074, respectively, Wilcoxon rank sum test with Holm’s correction for multiple comparisons), while the concomitant IgG response was equally strong in both groups (see. **Fig. S7A** for statistics). Clinically, mice of both groups reacted with an initial small increase in body temperature post challenge (a possible stress reaction measured also in unchallenged control mice) followed by a sudden drop in body temperature in mice challenged and sensitized using cholera toxin as adjuvant (**Fig. 9C**). The drop in body temperature was delayed, less pronounced, and recovered faster in *Cftr^-/-^* mice. Unchallenged mice or mice whose sensitization was without cholera toxin did not display signs of anaphylaxis (**Fig. 9D**), in contrast to peanut-sensitized and cholera toxin-treated *Cftr^-/ -^*mice and WT mice. In contrast, *Cftr^-/-^* displayed a delayed onset of anaphylactic reactions, a reduced peak, and a more pronounced recovery. Thus, *Cftr^-/-^* mice were partially protected from severe allergic reactions. Although the kinetic analyses showed a clear trend, the overall distributions between cholera toxin-induced *Cftr^-/-^* and WT mice did not significantly differ (**Figs. S7B** and **S7C**, p =0.415 and 0.0983, respectively, Wilcoxon rank sum test).

We then asked whether the cytokine profiles differed between the groups and whether they changed after challenge with peanuts. We speculated that a Th1 environment may skew toward productions of IgG, rather than IgE, which could alter the severity and course of an anaphylactic response. We chose three cytokines, IL-4, IL-10, and IFN-γ; the former a typical Th2 marker, the latter associated with a Th1-based inflammatory response. Naïve *Cftr^-/-^* and WT mice had no detectable IL-4 and IFN-γ levels in serum (with one exception) and median IL-10 levels of approximately 3.75 ng/ml (**Fig. 9E**). While serum IL-10 levels at week 5 and after challenge were close to baseline for both *Cftr^-/-^* and WT mice in the absence of cholera toxin treatment, cholera toxin-treated *Cftr^-/-^* and WT mice had IL-10 concentrations below detection. At week 5 pre-challenge, IL-4 and IFN-γ were slightly elevated in cholera toxin-treated *Cftr^-/-^* and WT mice compared with sensitized but non-cholera toxin-treated or non-peanut sensitized mice. Two hours post challenge, the IL-4 concentrations rose approximately 40-fold in peanut-sensitized cholera toxin-treated *Cftr^-/-^* and WT mice, whereas that of IFN-γ rose approximately 20-fold in peanut-sensitized cholera toxin-treated *Cftr^-/-^*, but not in WT mice.

In a small human cohort, plasma IFN-γ concentrations did not significantly differ between anti-Ara h2- seropositive individuals with CF (n=8), a collection of multimorbid hospital patients (including systemic lupus erythematosus, chronic kidney disease, and COVID-19, named ‘mixed’, n=26), and six healthy controls (**Fig. S7D**). However, plasma concentrations may only partially reflect local mucosal cytokine levels.

Together, there is a distinct cytokine signature in cholera toxin-treated *Cftr^-/-^* mice, visible post intraperitoneal (IP) challenge with peanut extract and characterised by high IFN-γ, low IL-10, and high IL-4. While IL-10 and IL-4 concentrations matched those in WT mice, high IFN-γ was a distinguishing feature at 5 weeks immunisation (p=0.0813, Wilcoxon rank sum test) and, most notably, after IP challenge (p=0.00117). This particular IFN-γ signature is remarkable as it has been shown to potently inhibit Th2 and Th17 cytokines (Sun et al., 2023), and its tolerogenic role has been described on few occasions (Ca, 2001; Noh & Lee, 2009).

## Discussion

We conducted a serological investigation to identify IgG against Ara-h2 in a large cohort of hospital patients and consulted the clinical records of individuals showing high titers to identify medical correlates of anti-peanut immunoreactivity. This approach relies on the availability and utilization of surplus samples collected for routine diagnostics, and of data associated with it. The study design is pragmatic (Ioannidis, 2016), it partially circumvents the usual enrolment bias (Grimes & Schulz, 2002), and enables the unconstrained generation of pathophysiological hypotheses. Our screen provided a strong indication that IgG against Ara-h2 are associated with CF. The most parsimonious explanation for this association would have been a higher prevalence of peanut allergy in pwCF, associated with IgE responses (Chen et al., 2021). However, this was not the case. We have therefore set to explore the immunological underpinnings of this association.

We identified high-affinity IgG antibodies against Ara-h2, but also against other peanut allergens and other food constituents, in the absence of IgE and without further indications that the repertoires of autoantibodies or alloantibodies in CF were globally perturbed. We found that such antibodies are affinity-matured due to food constituents rather than cross-reactivity with bacterial or viral proteins. We have additionally found that *CFTR* mutation carriers have the same prevalence of food allergies as pwCF, and these results are in agreement with observations in the population (Branum & Lukacs, 2009; Cianferoni & Spergel, 2009; Sicherer & Sampson, 2018; Soller et al., 2012; Tordesillas et al., 2017). Importantly, many of our findings have been reproduced in a cohort of paediatric pwCF from a different continent, bolstering our confidence in these observations. Lastly, we speculated that CF-associated gut pathology, in conjunction with a permissive cytokine environment, may be causative of the association; we therefore employed a *Cftr^-/-^* mouse model and found, upon allergic sensitisation with peanuts, a signature with low IgE and high IgG levels as well as an altered IFN-γ profile that associates with a milder anaphylactic reaction to peanut IP challenge when compared to WT mice.

Oral tolerance provides selective immunity to molecules ingested via the orogastric route in a microbially abundant environment. The IgG response to Ara-h2 was associated with pwCF, but with no other disease entity. While CF-related intestinal pathology is among the most burdensome aspects of the disorder, many other non-CF conditions also affect the gastrointestinal system. None of them displayed an association with IgG against Ara-h2, apart from what is mediated through CF. So how might this link with CF arise? CF is caused by the absence, deficiency or instability of the CFTR protein, an ion channel that maintains the balance of salt and water on many surfaces in the body. pwCF have a multiorgan manifestations leading to progressive lung disease, pancreatic insufficiency, CF-related diabetes, intestinal dysbiosis and inflammation, meconium ileus, intestinal obstruction, biliary cirrhosis and bronchiectasis (Filkins & O’Toole, 2015; Ooi & Durie, 2016). Moreover, there is strong evidence for an altered immune system in CF (Bojanowski et al., 2021; Bruscia & Bonfield, 2016, 2022; Hartl et al., 2012). In this regard, the absence of globally noteworthy aberrances at the level of the immunoglobulin repertoire in CF, shown in our study, might be surprising. We have further shown that CF is not associated with a higher prevalence of food allergies, both compared with *CFTR* mutation carriers and the general population. Consequently, there is an absence of a concomitant IgE response against Ara-h2 or other food ingredients, but an IgG response. The IgG response is dominated by IgG4, reminiscent of the profile observed in allergy patients after desensitization, i.e. following prolonged exposition to an antigen (Emmenegger et al., 2022; Irrgang et al., 2023; Pillai, 2023). Here, CD23^+^IgG1^+^ memory B cells poised to switch to IgE, upon activation with IL-4/IL-13, in food allergy (Jiménez-Saiz et al., 2017, 2018; Ota et al., 2023) appear to class-switch from IgG1 to IgG4, as in OIT (Jiménez-Saiz et al., 2018) and thereby lead to desensitization. Thus, one potential extrapolation from our observations could be a lower prevalence of peanut allergy in pwCF.

Exaggeration of defensive mechanisms to prevent ingestion of toxins can result in pathological food allergy (Florsheim et al., 2021). Therefore, to be able to elicit allergic sensitization, dietary proteins must be associated with noxious cues present in the food, as is shown e.g. with cholera toxin experiments performed in our study. A compromised epithelial barrier has been suggested to be involved in causing allergies (C. A. Akdis, 2021; Mattila et al., 2011). Because of lack of CFTR, epithelial integrity is compromised in the CF gut (De Lisle, 2014). Additionally, goblet cell hyperplasia is well-documented in CF (Garcia et al., 2009; Gustafsson et al., 2012; J. Liu et al., 2015) and goblet cells assume a pivotal role in supporting epithelial layer integrity and microbiota regulation (Gustafsson & Johansson, 2022; McDole et al., 2012; Weiner et al., 2011), with implications for oral tolerance (Gertie et al., 2022; Mazzini et al., 2014). In this context, pwCF may be more susceptible to epithelial malfunctioning, resulting in an overstimulation of the immune system by oral allergens.

It is extremely intriguing and unexpected that CFTR deficiency is associated with tolerogenic rather than anaphylactogenic responses to food allergens. Our peanut allergy sensitisation experiments conducted in wildtype and *Cftr^-/-^* mice indicated an altered serum cytokine profile, with an increase in IFN-γ concentration in CF. IFN-γ, unlike other IFN types, is selectively produced by T cells to inhibit Th2 T cell responses (Sun et al., 2023). Moreover, CF has also been associated with a predominant proinflammatory type 17 immunity (Price & O’Toole, 2021; Sheikh et al., 2023) and with increased IFN-γ concentrations in bronchoalveolar lavage fluid (Tiringer et al., 2013), while a Th2 T helper cell environment is predisposing to allergy (Abbas et al., 2018; Lloyd & Snelgrove, 2018; Tordesillas et al., 2017). However, the systemic cytokine signature does not necessarily reflect local mucosal milieus (Ferreira et al., 2009; Maina et al., 2022; Smith et al., 2021), and differences between the gut and the lung microenvironment are conceivable.

Based on our observations, the combination of a compromised gut epithelium, dysbiosis, and a proinflammatory cytokine milieu may (1) prime the immune system with antigens, in this case with a partially protease-resistant and heat-stable allergen, Ara-h2, in the proximity of noxious substances, and may then (2) poise the B cell response towards IgG4, a tolerance signal. Importantly, while colonization with *Pseudomonas* aeruginosa, which is common in pwCF, has shown to manipulate the host towards a type 2 immune response, including mucus production, to promote nutrient availability (Agaronyan et al., 2022), this immune bias may not generally apply to pwCF but perhaps only to a subset of them (Albon et al., 2023).

### Limitations of the study

This study analyzed a heterogeneous unselected cohort, a true excerpt of the population requiring medical aid in a university hospital setting. Its retrospective nature poses challenges owing to incompleteness of records and occasionally lack of relevant medical details. We have only restricted knowledge of whether certain pwCF had received mutation-specific treatment with potentiators like Ivacaftor (Swissmedic approval: 2012) or combinations of Ivacaftor with correctors like Lumacaftor (Swissmedic approval: 2016) or Elaxacaftor and Tezacaftor (Swissmedic approval: 2020), although the time of sampling (75% until end of 2017, 21% in 2018, less than 4% in 2019) suggests that this treatment had not been employed widely. Moreover, the specific mutation was not always available for all pwCF. Similar limitations pertain to the availability of detailed information on patients’ allergies in the medical records. Allergies are often selfreported, without being serologically and/or clinically confirmed, and were interpreted with due caution. To offset these limitations, we have utilized complementary methodologies, we have examined multiple cohorts to validate findings, and experiments were conducted at multiple sites. Finally, we have translated an observation on humans onto a model system, a *Cftr^-/-^* mouse. A particularly intriguing prediction derived from the above findings is that transient CFTR inhibition, e.g. in antigen-presenting cells like macrophages or B lymphocytes expressing CFTR (Bruscia & Bonfield, 2022), may quench allergogenic immune responses and might become useful in the context of desensitisation regimens.

## STAR★Methods

### Key resources table

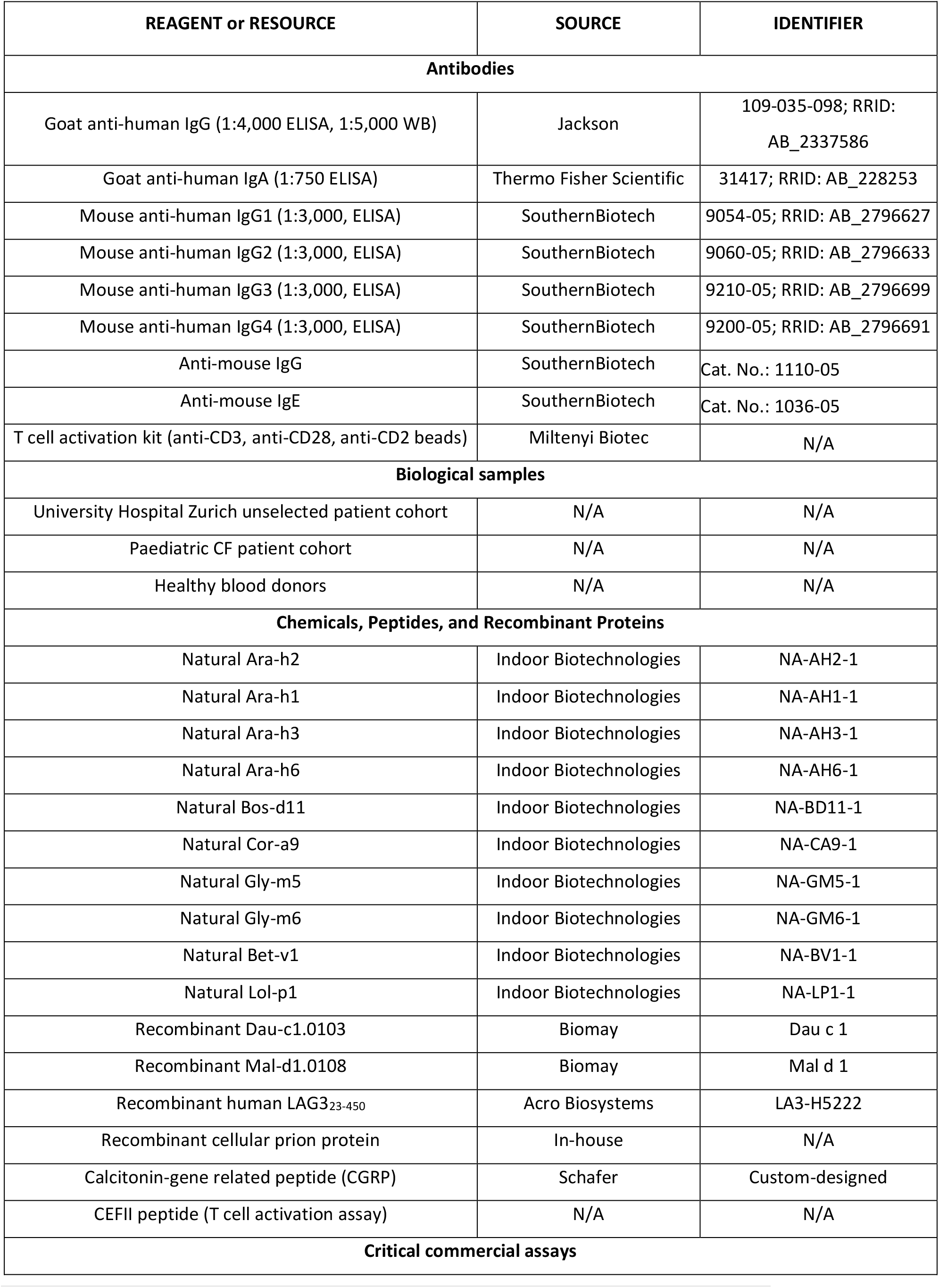
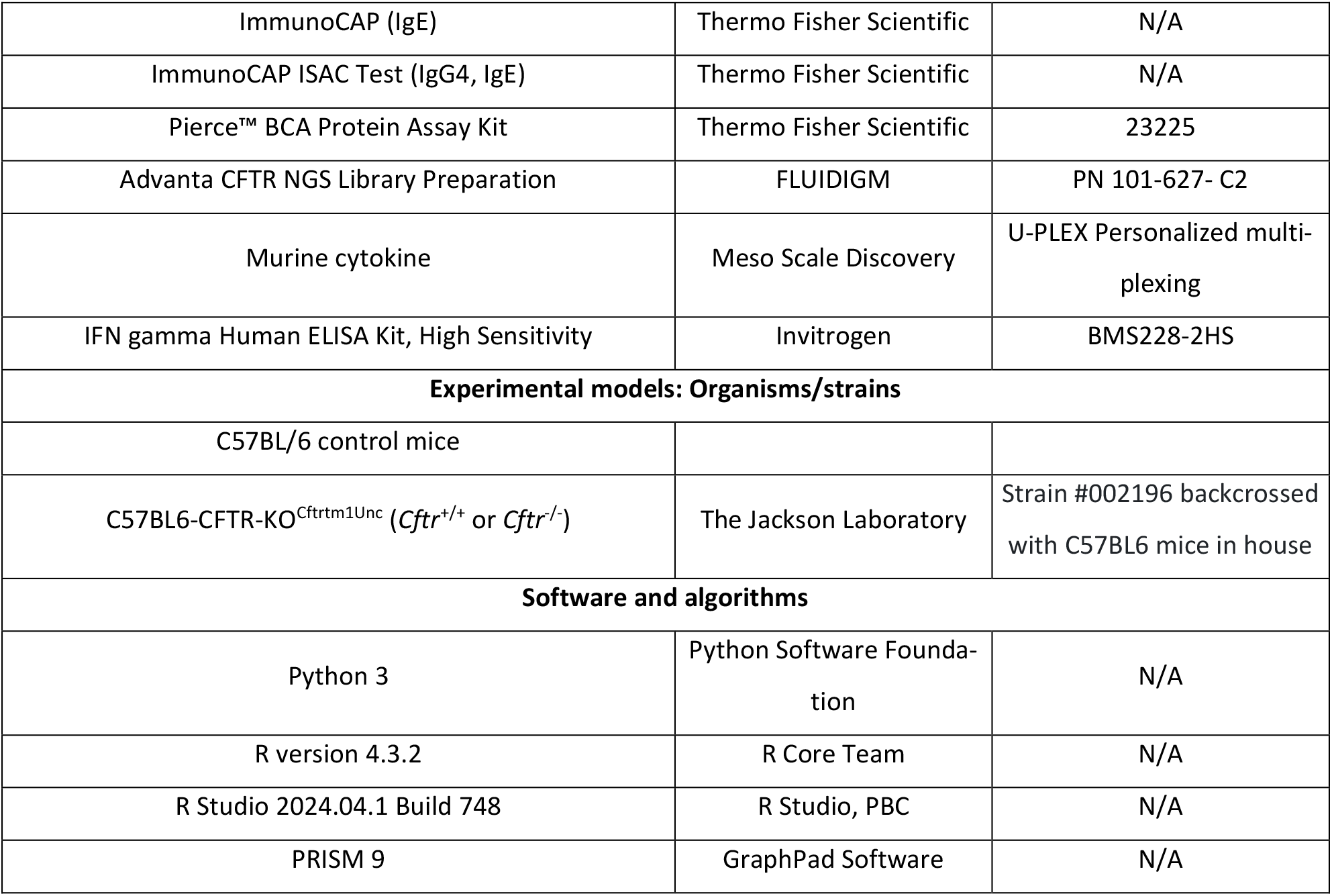

### Resource Availability

#### Lead contact

Further information and requests for resources should be directed to and will be fulfilled by the lead contact, Marc Emmenegger.

#### Materials availability

Small amounts of the biological samples can be shared if available, upon reasonable request, and if approval by an ethics committee as well as an MTA is in place. This study did not generate new unique reagents.

### Data and code availability

- Specific data sets can be shared upon reasonable request and if approval by an ethics committee as well as a data transfer agreement is in place.
- Code employed in this study will be publicly available on Zenodo in the peer-reviewed version of this manuscript after publication.
- Any additional information required to reanalyse the data reported in this paper is available from the lead contact upon request.

### Experimental model and subject details

#### Ethics statement

All experiments and analyses involving samples from human donors were conducted with the approval of the local ethics committee (*Ethikkommission Kanton Zürich*, KEK-ZH-Nr. 2015-0561, BASEC-Nr. 2018- 01042, and BASEC-Nr. 2020-01731, Yale IRB #0906005332) and were approved by the University of Zurich, the University Hospital Zurich, and the Yale University Medical School Human Investigation Committee, in accordance with the provisions of the Declaration of Helsinki and the Good Clinical Practice guidelines of the International Conference on Harmonisation. Part of the study conducted on anonymized data was performed after clarification of responsibility where a declaration of no-objection was obtained (*Ethikkommission Kanton Zürich*, Req-2019-00572 and Req-2022-00638). Whenever required, written general or informed consent was obtained from study participants.

#### Study design and sampling

To study the prevalence of IgG antibodies against nAra-h2, we made use of surplus plasma samples (n=27,239) from inpatients and outpatients admitted to the University Hospital of Zurich (USZ, n=24,536) collected daily (Monday-Friday) and used for population-wide interrogations of the antibody repertoire (Emmenegger, De Cecco, et al., 2023; Magalhães et al., 2021; Senatore et al., 2020). The criteria for our study to include a sample into the analysis were: (1) The patients’ blood was sent to the Institute of Clinical Chemistry (at USZ), (2) there was enough residual heparin plasma (150 µl) for the automated generation of a research aliquot, (3) no aliquot from the same patient was already provided within the same month, (4) additional information (age, sex, clinical ward to which patient was admitted) was available, as described recently (Emmenegger, De Cecco, et al., 2023). The demographic characteristics of this patient collective is described in **Table 1**. Briefly, our unselected patient cohort comprised 48% female (n=11,944) and 52% male (n=12,592) patients, with a median age of 54 (IQR: 49-68) years.

For secondary screens and as controls, we additionally investigated samples from blood donors of the Blood Donation Service of the Canton of Zurich who consented to further use of their samples for research. This collective was characterized by 40% male and 60% sex participants and a median age of 44 (IQR: 31-55) years. The criteria to be admitted for blood donation are in line with international standards of blood donation services, see (Blutspendedienst, 2021a) and are as outlined before (Emmenegger, De Cecco, et al., 2023). The detailed inclusion and exclusion criteria are enumerated here (Blutspendedienst, 2021b). Moreover, blood samples from children with CF were obtained during routine clinic visits at the Yale Paediatric CF Center with informed consent in accordance with relevant laws and institutional guidelines. This cohort consisted of 45% female and 55% male participants with a median age of 13 (9-17) years, as outlined in **Table 1**.

151 individuals participated in an online health survey. Their median age was 36 (IQR:16.5-43) years. 62.3% of participants were female and 37.7% male (see **Table 3**). Additionally, a registry-based study was conducted in 115 individuals (median age 5.0 (IQR: 2.0-15.0), 48.7% of female sex) whereof 56 individuals were *CFTR* mutation carriers, and 59 individuals were pwCF (see **Table 4**).

### Method Details

#### Exploratory high-throughput screening

The profiling of antibodies in an unselected university hospital patient cohort using a custom antibody discovery platform was conducted as recently shown (Emmenegger, De Cecco, et al., 2023; Emmenegger, Kumar, et al., 2021; Senatore et al., 2020), with minor modifications. Heparin plasma samples (hospital patients) or EDTA plasma samples (healthy blood donors) were investigated for the presence of antibodies directed against the major peanut allergen, natural Arachis hypogaea 2 (nAra-h2) purified from peanut extract. As control, some samples were tested for the presence of antibodies against calcitonin-gene related peptide (CGRP). We first coated high-binding 1536-well assay plates (Perkin Elmer SpectraPlates) with 0.5 µg/ml antigen, at a volume of three µl/well (end volume in all subsequent dispensing steps, except for blocking) on the Fritz Gyger CertusFlex or the Thermo Fisher Multidrop Combinl nanodispenser. Plates were incubated for one hour at 37 °C in an automated Thermo Fisher plate incubator and subsequently washed three times with PBS 0.1% Tween-20 (wash buffer) on the Biotek El406 washer/dispenser. Blocking with 10 µl/well 5% milk in PBS 0.1% Tween-20 (blocking buffer) was performed on Biotek MultifloFX using the peristaltic dispensing unit. Following the incubation at RT for one hour and the subsequent aspiration of blocking buffer using Biotek El406, plates were sealed with Agilent plate sealer and stored at -30 or -80°C until the day of experiment. Frozen plates were thawed and peeled with Brooks plate peeler. 1536-well assay plates were pre-filled with 1% milk in PBS 0.1% Tween-20 (sample buffer) according to a pre-defined plate layout using Fritz Gyger CertusFlex or Thermo Fisher Multidrop Combinl, and plates entered the ECHO555 acoustic dispenser (Labcyte) where pre-diluted plasma samples were dispensed at different volumes (from 1200-7.5 nl) from a source plate using ultrasound. Reaching a final well volume of three µl, the samples were thus diluted between 0.02 and 1.67x10^-4^. Plates were incubated for two hours at RT and washed five times with wash buffer. Secondary antibody (Peroxidase AffiniPure Goat Anti-Human IgG, Fcγ Fragment Specific, Jackson, 109-035-098, at 1:4,000 dilution in sample buffer) was dispensed on the MultifloFX peristaltic pump, incubated for one hour at RT, and washed three times. Lastly, TMB was added, incubated for three minutes at RT, and the chromogenic reaction was stopped with 0.5 M H_2_SO_4_. The plates were centrifuged after all dispensing steps, except for the addition of TMB. The absorbance at 450 nm was measured in a plate reader (Perkin Elmer, EnVision). Optical densities were fitted using logistic regression as recently introduced (Emmenegger, De Cecco, et al., 2023). To filter out potential false-positive results from the HTS due to outliers, the quality of the regression was assessed by means of the mean squared residual error. Thus, only plasma samples with a p(EC_50_) ≥ 2 and the mean squared residual error of the logistic regression < 20% of the actual p(EC_50_) value were considered as hits, an approach we have applied before (Senatore et al., 2020).

### Isolation of peripheral blood mononuclear cells and plasma from patients

Peripheral blood mononuclear cells (PBMCs) were isolated from patients as described (Emmenegger, De Cecco, et al., 2021). Briefly, patients were re-contacted via the Clinical Trial Center at University Hospital Zurich and consented for a blood donation. The blood was first diluted 1:2 with sterile PBS, separated via Ficoll gradient, purified using 10% B cell medium (IMDM (21980-065, Gibco), 10% FBS, 1x P/S (15140-122, 200x stock, Gibco), 1x MEM non-essential amino acids (11140-035, 100x stock, Gibco), 1x MEM sodium pyruvate (11360-039, 100x stock, Gibco), and β-mercaptoethanol (1:1,000, 31350-010, Gibco)) diluted in PBS, and aliquots of 5-10 million PBMCs/cryo vial were frozen in FBS with 10% DMSO. The 1:2 diluted plasma was harvested and used for experiments.

### B cell receptor repertoire analysis

High quality RNA (RIN>7) was extracted from total PBMCs and reverse-transcribed into cDNA with an oligo dT primer. A unique molecular identifier (UMI) and an adaptor sequence were added to the cDNA prior to purification with streptavidin magnetic beads. VDJ amplification of IGHA, IGHD, IGHE, IGHG, IGHM genes and index barcode incorporation was done with a primary PCR reaction (PCR1). PCR products were purified with AMPure XP beads and subjected to a secondary PCR reaction (PCR2) whereby dual-indexed primers (i7 and i5) were added along with the Illumina P5 adaptor. A qPCR reaction, prior to PCR2, was used to determine the optimal number of cycles to use to avoid the plateau phase. PCR2 products were purified and measured on an Agilent Tapestation. Samples were pooled equimolarly and 20% PhiX sequences were added to the pool. The pool was sequenced on the Illumina MiSeq platform in 2x300 bp paired-end, sequencing mode. For the analysis of the read data we relied on the Immcantation suite Version 4.0.0 (https://immcantation.readthedocs.io). The analysis steps followed the workflow described in (Vander Heiden et al., 2014) and relied on the tools presto 0.6.0, changeo 1.0.0, alakazam 1.0.1, shazam 1.0.0 and tigger 1.0.0.

We applied the manual pRESTO pipeline to perform quality control, filter and process the reads, as described (Vander Heiden et al., 2014). As an output we generated consensus sequences and the assembled receptor sequences. In a modification of the workflow, we performed deduplication of the UMIs with the package UMI-Tools (https://github.com/CGATOxford/UMI-tools) and allowed two mismatches when clustering UMIs.

For the subsequent postprocessing and analysis of the clone sequences, we used the pipeline templates provided in the Immcantation tools and custom R scripts to perform the following steps: performs V(D)J <u>alignment using IgBLAST and post-processes the output into the Change-O data standard (changeo-igblast</u> pipeline), infer V segment genotypes using TIgGER (tigger-genotype pipeline), automated detection of the clonal assignment threshold (shazam-threshold pipeline), assign Ig sequences into clonally related lineages and build full germline sequences (changeo-clone pipeline). All pipelines were run with default settings. With the corresponding R packages (tigger and shazam), we estimated the abundances of the BCR clones, and we computed the Hill diversity curves (alakazaam package). Somatic hypermutation and mutational frequencies were assessed with the shazam package.

### Analysis of convergence between sequences obtained through BCR repertoire sequencing and publicly available repertoires

Heavy and light CDR3 sequences were mapped to publicly available CDR3 amino acid sequence datasets (Aguzzi et al., 2020; Croote et al., 2018; Hoh et al., 2016) with a 90% identity using the usearch tool (Version 11.0.667). The usearch performs a local alignment and we required a coverage of at least 70% of the query sequences.

### HLA genotyping, epitope prediction, and T cell activation assay

Genomic DNA extracted from the peripheral blood of six CF patient samples was typed for HLA-DRB1, - DRB345, -DQA1, -DQB1, -DPB1, and -DPA1 alleles by 1x resolution typing NGS (HistoGenetics, Ossining, NY USA). We compared the observed allele frequencies using the following database: http://allelefrequencies.net/hla.asp, (Gonzalez-Galarza et al., 2020). The alignment and comparison of the amino acid sequences of HLA-DQA1*05:01:01 with the DQA1*01:01:01 allele and other DQA1* alleles of the six individuals were performed using https://www.ebi.ac.uk/ipd/imgt/hla/alignment/, (Barker et al., 2023). In silico peptide binding predictions of Ara-h2 to the HLA-DQA/B combinations DQA1*05:02/DQB1*02:01:01 and DQA1*05:02/DQB1*03:01:01 were performed using NetMHCII-pan-4.0; see (Jensen et al., 2018). Prediction values are given in nM IC50, calculated as a % rank to a set of 1,000,000 random natural peptides, and indicated as strong (≤ 2%) or weak (≤ 10%) binding peptides. The following parameters were used for the algorithm: threshold: -99.9; threshold for strong binder (% Rank): 2; Threshold for weak binder (% Rank): 10. The sequence of Ara-h2 used was (Uniprot-ID: Q6PSU2): MAKLTILVA LALFLLAAHA SARQQWELQG DRRCQSQLER ANLRPCEQHL MQKIQRDEDS YGRDPYSPSQ DPYSPSQDPD RRDPYSPSPY DRRGAGSSQH QERCCNELNE FENNQRCMCE ALQQIMENQS DRLQGRQQEQ QFKRELRNLP QQCGLRAPQR CDLEVESGGR DRY.

### Sequencing of the CFTR gene in patients with peanut allergy

DNA from PBMCs of an anonymised collection of 90 confirmed patients with allergies and of four pwCF was extracted using the Qiagen DNeasy Blood & Tissue kit. We then followed the *Advanta CFTR NGS Library Preparation on the LP48.48 IFC with Juno* protocol (FLUIDIGM, PN 101-627- C2 PROTOCOL, Fluidigm Corporation, South San Francisco, California, United States) to create suitable libraries for subsequent amplicon sequencing. We aimed to perform a target enrichment of *CFTR* variants from each of the 27 exons and select intronic regions. In short, we first pre-amplified the genomic (g)DNA as the concentration after gDNA purification was < 30 ng/µl. The pre-amplified gDNA samples (n=96 samples, 90 samples from patients with allergies, 4 samples from PwCF as positive controls, and 2 H_2_O samples as assay negative controls) were then indexed in a 96-well plate using targeted DNA sequencing barcodes, mixed with sample pre-mix (PCR water, TSP master mix, TSP sample loading reagent, TSP DNA polymerase), and were loaded into a flow cell containing integrated flow circuits (IFCs), together with the *CFTR* assay mix (loading agent and *CFTR* NGS assays pools) and the non-assay mix (loading agent with *CFTR* NGS assay pools). The JUNO scripts were run according to the manufacturer’s instructions and the pooled samples were cleaned up following the protocol. This process was run twice for 48 samples per round as the IFCs used contained 48 wells, and we obtained pools of 48 uniquely barcoded samples twice, i.e. 96 samples in total. Next, the sequencing adapter was added to the purified pooled libraries, another PCR was run as detailed by the manufacturer, and the PCR products were cleaned up. The PCR products after the first PCR round (PCR1) and after the second PCR round (PCR2) were then quality controlled on a Qubit Fluorometer (Thermo Fisher) following the manufacturer’s instructions. We obtained the following concentrations: for library 1, after PCR1: 3.96 ng/µl; for library 2, after PCR1: 2.47 ng/µl; for library 1, after PCR2: 10.9 ng/µl; for library 2, after PCR2: 9.15 ng/µl. We then looked at the samples with the TapeStation (Agilent) on an Agilent High Sensitivity DNA chip and observed the expected right shift of the electropherogram with a peak between 300-400 bp. The library preparation was performed at the Genomic Diversity Centre, ETH Zurich. For Illumina sequencing, the two libraries were pooled and paired end 150 bp sequencing was performed on an Illumina MiSeq system at the Functional Genomics Centre Zurich (FGCZ). The quality of the raw data was assessed with FastQC 0.11.9. The reads were subsequently filtered with fastp 0.20.0 for minimum length of 18bp and mapped to the GRCh38 reference genome with Bowtie2 2.4.1. Marking of duplicates was performed with Picard tools 2.22.8 and sorting and indexing of the alignment was performed with samtools 1.11. Qualimap 2.2.1, SAMstat 1.5.1 and Picard tools 2.22.8 were used for assessing the quality of the mapping. Single nucleotide variants were called using GATK “HaplotypeCaller” pipeline (4.2.0) first for each patient data separately and then joint genotyping across the whole cohort was performed with GATK “CombineGVCFs” and “GenotypeGVCFs” pipelines (4.2.0), as recommended by the GATK Best Practices. Vcftools 0.1.16 and bcftools 1.9 were used to calculate allele frequency and proportion of heterozygosity for each variant across the 90 Peanut-allergic patients, excluding the 4 CF control patients. Variants of low coverage (DP < 200) and detected outside of the *CFTR* gene were discarded with SnpSift 4.3. The identified sequence variants within the *CFTR* gene were then functionally annotated using SNPeff 4.3. Additional annotations were added with ClinVar (downloaded on the 07.03.2022) for a better understanding of the general clinical significance, with gnomad v3.1.1 allele frequencies using the pipeline available at https://github.com/KalinNonchev/gnomAD_DB and with data from CFTR2 database (https://cftr2.org/, 29.04.2022). The graphs were created in *Python 3*.8.3.

### Online health survey to correlate health status with CFTR genotype

We performed a fully anonymised data collection and data interpretation to correlate the *CFTR* genotype with health. The aim of the questionnaire was to identify whether the presence or absence of mutations in the *CFTR* gene were associated with allergies, in particular in heterozygous mutation carriers. We first designed a survey using REDCap (see **Supplemental File 13** for survey design and survey questions), ensuring that no clear link to allergies was visible to prevent bias. The online health survey was then announced on online platforms of CF patient organisations, notably Mukoviszidose e.V. Everybody whose *CFTR* genotype was known or was the parent of a child with CF, was amenable to participate. Data was analysed in R, similarly to previous accounts (Emmenegger, De Cecco, et al., 2023).

### Registry-based and diagnostic code-based inference of prevalence of food allergy in mutation carriers and in pwCF

PwCF (those confirmed by genetic sequencing of the *CFTR* gene) were identified using the Yale Paediatric CF Foundation’s Patient Registry. This is a voluntary registry that captures the demographics and health information of pwCF who receive care in CF Foundation-accredited care centres in the United States. In parallel, *CFTR* mutation carriers (those with International Classification of Diseases, Tenth Revision (ICD-10) codes of Z14.1) were identified using Yale New Haven Health System’s electronic medical records. Only *CFTR* mutation carriers with at least one clinic encounter at Yale from March 2018 to March 2023 who were between the ages of 0-18 at the time of that visit were included in the analysis. Data was analysed in R, similarly to previous accounts (Emmenegger, De Cecco, et al., 2023).

### High-throughput microELISA of pwCF and controls using antigen panel

The ELISA was conducted as detailed above (see Exploratory high-throughput screening), with the following modifications. (1) Instead of the unselected patient cohort, PwCF were selected from the biobank and random patient controls and healthy blood donor controls were included. (2) Instead of merely coating with nAra 2, an antigen panel was used (see list of antigens).

### Indirect ELISA for IgG, IgG1, IgG2, IgG3, IgG4, and IgA with plasma samples

Indirect ELISA was conducted in 384-well high-binding plates (PerkinElmer SpectraPlate) as previously shown. In short, plates were coated with 0.5 ug/ml nAra-h2 in sterile PBS and incubated at 37 °C for 1 h, followed by 3 washes with PBS 0.1% Tween-20 using Biotek El406 and by blocking with 40 µL 5% milk in PBS-T for 1.5 h. Serum samples were diluted in sample buffer (1% milk in PBS 0.1% Tween-20) and a serial dilution was carried out (volume: 20 µL/well). After the sample incubation for 2 h at RT, the wells were washed five times with wash buffer and the presence of IgG, IgG1, IgG2, IgG3, IgG4, IgA, antibodies directed against nAra-h2 was detected using a panel of secondary antibodies, validated previously (Emmenegger et al., 2022) and referred to in **Key Resource Table**. The incubation of the secondary antibodies for one hour at RT was followed by three washes with PBS 0.1% Tween-20, the addition of TMB, an incubation of five minutes at RT, and the addition of 0.5 M H_2_SO_4_. The plates were centrifuged after all dispensing steps, except for the addition of TMB. The absorbance at 450 nm was measured in a plate reader (Perkin Elmer, EnVision).

### ISAC and ImmunoCAP

IgG4 and IgE ISAC (Thermo Fisher Science) were carried out according to the manufacturer’s guidelines, using an array of 112 allergens from 48 sources, including food (oral), pollen (airway), venom, and contact allergens (Matricardi et al., 2016). ImmunoCAP (Thermo Fisher Science) assays were performed according to the manufacturer’s guidelines.

### Rapid Extracellular Antigen Profiling

REAP was performed as previously described (Wang, Dai, et al., 2021). In brief, IgG was purified from 50 µl serum by 20 µl protein G magnetic resin (Lytic Solutions) and quantified by NanoDrop 8000 Spectrophotometer (Thermo Fisher Scientific). Purified IgG was incubated with 10 OD induced empty yeast (same strain as the one used in library but displaying empty vector) for 3 hours in 4 °C with shaking to remove unspecific yeast binding IgG, and yeast-depleted IgG was filtered with 96-well 0.45-μm filter plates (Thomas Scientific). 10 μg yeast-depleted IgG was incubated with 10 OD induced library yeast in 100 µl PBE (PBS with 0.5% BSA and 0.5 mM EDTA) with shaking for 1 hr at 4 °C, followed by 30 mins incubation with 1:100 biotin anti-human IgG Fc antibody (clone HP6017, BioLegend) in 100 µl PBE and 30 mins incubation with 1:20 Streptavidin MicroBeads (Miltenyi Biotec) in 100 µl PBE. Streptavidin MicroBeads captured yeast was positively selected by Multi-96 Columns (Miltenyi Biotec) placed in MultiMACS M96 Separator (Miltenyi Biotec). Selected yeast were recovered in 1 ml SDO −Ura at 30 °C for 24 hrs. DNA was extracted from yeast libraries using Zymoprep-96 Yeast Plasmid Miniprep kits (Zymo Research). Purified plasmids were amplified and indexed (2 rounds Phusion PCR, 24 cycles/round), gel purified, and sequenced on an Illumina MiSeq Instrument with MiSeq Reagent Kit v3 (150-cycle). REAP score was calculated as previously described (Wang, Dai, et al., 2021). In brief, it is the log2-fold change between the frequency of each protein in pre and post selection library, i.e. before and after addition of the patient-derived purified IgG antibodies.

### Competitive Western Blotting

Strips for competitive Western Blotting were prepared as follows: 100 ng nAra-h2 and 100 ng BSA were loaded into the same lane of NU-PAGE 4-12% Bis-Tris gel (ThermoFisher), run at 80 V for 10 min and then at 120 V for 60 min in MES running buffer. Gels were transferred onto PVDF membranes with a dry transfer system (iBlot 2 Gel Transfer Device, ThermoFisher) at 20 V for 7 min, cut into strips, and the membranes were blocked with 5% SureBlock (Lubio Science) for 1 hour at RT. Next, 1:100 dilutions of plasma samples were prepared in 1% SureBlock in PBS 0.1% Tween-20 (sample buffer) and matched samples were (i) mixed with soluble 1 µg/ml nAra-h2 (competition) or (ii) without addition and incubated for two hours at 4° C in Eppendorf tubes. The pre-incubated plasma samples (both (i) and (ii)) were then added to the PVDF membrane strips and incubated overnight at 4° C. The following day, membranes were washed four times with PBS 0.1% Tween-20 and incubated for one hour with HRP-linked anti-human IgG antibody (Peroxidase AffiniPure Goat Anti-Human IgG, Fcγ Fragment Specific, Jackson, 109-035-098, at 1:5,000 dilution in sample buffer) at RT. Following another four-times wash with wash buffer, Immobilon Crescendo HRP Substrate (Merck Millipore) was added to the strips and the membranes were immediately imaged on the Fusion SOLO S imaging system (Vilber).

### Purification of IgG fraction from patient plasma

10 mL 1:2 pre-diluted patient plasma (i.e. 5 mL undiluted plasma) collected during the isolation of peripheral blood mononuclear cells (PBMCS) (Emmenegger, De Cecco, et al., 2021) was first filtered through a sterile 0.4 µM filter. 2.5 mL Protein G Sepharose 4 fast flow (17-0618-05) was loaded onto a liquid chromatography column (Luer Lock, non-jacketed, I.D. x L 1.0 cm x 10 cm, Sigma), washed with 15-20 mL dH_2_O, and then equilibrated with 15-20 mL PBS. The pre-filtered plasma samples were then slowly loaded to the column, and the column was washed with 15 mL PBS. The tubing of the ÄKTA was then washed with dH20 and equilibrated with 0.1 M Glycine PH 2.7. The IgG fraction bound to Protein G on the column was then eluted using 20 mL 0.1 M Glycine pH 2.5 at a flow rate of 2.5 mL/min. The eluates were collected in tubes containing 50 µL 1 M TRIS pH 8, pooled, and dialysed against PBS overnight at 4 °C, resulting in 20 mL purified IgG per sample. We thus recovered 14-23 mg total IgG per patient, at total IgG concentrations between 0.7-1.15 mg/mL. The total IgG concentration in undiluted plasma is approximately four times higher, as the undiluted plasma (input: 5 mL) was diluted four times (output: 20 mL).

### Affinity determination using microfluidics diffusional sizing

Alexa Fluor 647 N-hydroxysuccinimide ester (in DMSO, 4 equivalents) was added to nAra-h2 (typically 3.88 nmol, 1 equivalent) in 0.1 M NaHCO3 (pH = 8.2). The mixture was incubated for 1 h at ca. 20 °C, protected from light, and subsequently purified by size exclusion chromatography on a Superdex 200 increase column connected to an ÄKTA pure instrument (GE Healthcare, UK) with a flow rate of 0.5 mL/min and PBS as eluent buffer, to yield labelled nAra-h2 (1.37 nmol).

Labelled HLA was incubated with varying fraction of purified IgGs derived from patient plasma and diluted in PBS 0.1% Tween-20 (Sigma Aldrich, US). The samples were incubated at RT for 30 minutes and the hydrodynamic radius, Rh, was determined by microfluidic antibody affinity profiling on a Fluidity One W (Fluidic Analytics).

The dissociation constant, 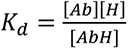, and the antibody binding sites concentration, [Ab]_tot_, was determined through Bayesian inference, as previously described (Fiedler et al., 2021; Schneider et al., 2022,

2023). [Ab] and [R] are the equilibrium concentrations of antibody binding sites and nAra-h2, respectively, and [AbR] is the concentration of bound nAra-h2. The data were analysed by Bayesian inference, according to the following equations. Following correction of fluorescence intensities for plasma autofluorescence, the fraction, *f*_*d*_ of nAra-h2 to diffuse into the distal channel is defined by (Linse et al., 2020)

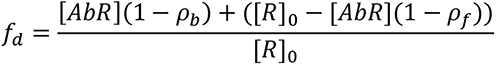

where [*R*]_0_ is the total concentration of nAra-h2, and *ρ*_*b*_ and *ρ*_*f*_ are the fractions of bound and free nAra- h2 to diffuse into the distal channel, respectively. By solving the binding equation, we obtain the following expression for [*AbR*]

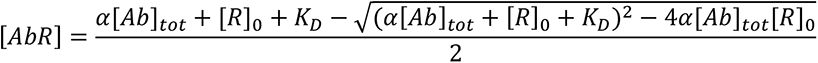

where α is the fraction of plasma used in the measurement and [*Ab*]_*tot*_ is the total concentration of antibody binding sites in the sample. K_d_ and [*Ab*]*tot*were thus determined through Bayesian inference, with *ρ*_*b*_ and *ρ*_*f*_ as additional parameters to be inferred. The prior was considered to be flat in logarithmic space for K_d_ and [*Ab*]*tot*, flat in linear space for *ρb* and *ρf*. The likelihood function was considered to be Gaussian, with a standard deviation obtained through triplicate measurements.

### IFN-γ measurement in human plasma

Measurement of IFN-γ was performed according to the manufacturer’s guidelines using the IFN gamma Human ELISA Kit, High Sensitivity (Invitrogen, BMS228-2HS). In short, 1:2 pre-diluted plasma samples (with PBS) were assayed in duplicates. The resulting OD was then interpolated with GraphPad Prism using a Sigmoidal, 4PL, X is log(Concentration) function. The resulting interpolated concentration was multiplied by four to account for the 1:2 predilution and the 1:2 dilution performed during the assay, and the concentration in pg/ml was calculated.

### Allergic sensitisation and challenge of Cftr^-/-^ and wild type mice

Transgenic CFTR^−/+^ (B6.129P2-KOCftrtm1UNC; CF) mice were originally purchased from the Jackson Laboratory and bred with C57BL6 mice in the Yale animal facilities for over 5 generations. Cohoused, age (16-20 week-old mice) - and sex-matched *Cftr^-/-^* and *Cftr^+/+^* (WT) were sensitized to peanut allergen weekly for 6 weeks as previously described (Gertie et al., 2022). Briefly, a freshly prepared 5 mg ground blanched peanut was dissolved in 200 µl of 0.2 mol sodium bicarbonate buffer with or without 10 µg of cholera toxin (List Biologicals, Campbell, Calif) as an adjuvant and delivered by oral gavage. Blood was collected from the retro-orbital plexus from day 14 every 2 weeks until the endpoint, and plasma was isolated and preserved at -80 °C. After the 6 weeks of sensitization, mice were permitted to rest for 10 days before the challenge. Mice received no food for 4 hours before being challenged intraperitoneally with 500 μg crude peanut extract (Greer, Cambridge, Mass) in 200 μL PBS. Rectal temperature was measured every 15 minutes for up to 60 minutes, clinical symptoms scores were assessed every 15 minutes (as described in **Fig. 9D**), and sera were collected for analysis 1 hour after the challenge. All procedures were performed in compliance with relevant laws and institutional guidelines and were approved by Yale University Institutional Animal Care and Use Committee.

### Cytokine measurement in murine plasma

The cytokine concentrations in the plasma of *Cftr^-/-^* and WT mice were assessed with a U-plex Meso-Scale Multiplex immunoassay platform following the manufacturer’s instructions (*Meso Scale Diagnostics, LLC). The following cytokines were simultaneously evaluated in 50 μl of* plasma: IFN-γ, IL-10 and IL-4.

### IgG and IgE ELISA with murine plasma

Serum samples were analysed by ELISA to measure peanut-specific antibodies. Briefly, 20 μg/mL of crude peanut extract (Greer Laboratories) in carbonate-bicarbonate buffer (pH 9.6) was coated on 96-well Maxisorp plates (Thermo Fisher Scientific) overnight at 4 °C. Plates were blocked with 1% bovine serum albumin (BSA) in PBS at RT for 1-hour followed by the addition of serially diluted serum samples with a 2-hour incubation at 37 °C. Peanut-specific antibodies of each isotype were detected with horseradish peroxidase (HRP)-conjugated goat anti-mouse IgE and, goat anti-mouse IgG , goat anti-mouse IgG1(Polyclonal, Southern Biotech) antibodies at 37 °C. Antigen (Ag)-specific antibodies of each isotype were obtained from pooled sera from repeatedly immunized mice and used as standards, represented as arbitrary units (AU)/mL, for Ag-specific ELISAs.

### Identification of homologous sequences

A protein BLAST was conducted using the BLOSUM-62 matrix on UniProt (Altschul et al., 1997; Schäffer et al., 2001). We have applied an E-Threshold of 0.01 without any further filtering, gapped=yes, and queried the UniProtKB reference proteomes plus Swiss-Prot (uniprotkb_refprotswissprot, 64,899,224 sequences; 25,452,742,688 total letters). The sequence of Ara-h2 used was (Uniprot-ID: Q6PSU2): MAKLTILVA LALFLLAAHA SARQQWELQG DRRCQSQLER ANLRPCEQHL MQKIQRDEDS YGRDPYSPSQ DPYSPSQDPD RRDPYSPSPY DRRGAGSSQH QERCCNELNE FENNQRCMCE ALQQIMENQS DRLQGRQQEQ QFKRELRNLP QQCGLRAPQR CDLEVESGGR DRY.

### Quantification and statistical analysis

#### Exploratory data analysis

PCA and UMAP were conducted as shown before (Emmenegger, De Cecco, et al., 2023). In short, PCA was carried out using the default implementation in the R stats package (prcomp, https://www.rdocumentation.org/packages/stats/versions/3.6.2/topics/prcomp) and data was visualized using the factoextra package (https://cran.r-project.org/web/packages/factoextra/index.html). For UMAP, the following configuration parameters from the umap package in R (https://CRAN.R-project.org/package=umap) were used, all of which are default, except for the metric where cosine was sometimes used instead of Euclidean due to the binary nature of the data (n_neighbors: 15, n_components: 2, metric: cosine, n_epochs: 200, in-put: data, init: spectral, min_dist: 0.1, set_op_mix_ratio: 1, local_connectivity: 1, bandwidth: 1, alpha: 1, gamma: 1, negative_sample_rate: 5, a: NA, b: NA, spread: 1, random_state: NA, transform_state: NA, knn: NA, knn_repeats: 1, verbose: FALSE, umap_learn_args: NA). UMAP data was plotted using ggplot2 in R. Hierarchical clustering was conducted using the stats::hclust (https://www.rdocumentation.org/pack-ages/fastcluster/versions/1.2.3/topics/hclust) library and dendrograms were visualized with functions of dendextend (https://cran.r-project.org/web/packages/dendextend/vignettes/dendextend.html), circlize (https://jokergoo.github.io/circlize_book/book/), and factoextra. The method for hierarchical clustering was ‘binary’ when using the absence or presence of an ICD-10 code as feature or ‘ward.D2’ when using p(EC_50_) values.

### Statistical data analysis

Using a reactivity threshold of p(EC_50_) [sample dilution] ≥ 2 to define the seropositive samples (‘hits’ or ‘seropositives’) exceeding the threshold and the seronegative samples (‘non-hits’ or ‘seronegatives’) with p(EC_50_) values < 2, we aimed to correlate ICD-10 codes, active ingredients of prescribed drugs, and sex (male/female) with seropositivity. To this end, we first aggregated the dataset to obtain medical data occurring at least 5 times (i.e. in > 3.75%) in the seropositive group and then conducted χ^2^ statistical testing, at *α-level* 0.01 and applying Bonferroni correction for multiple comparisons. Χ^2^ analysis was performed using the stats.chi2_contingency function of the SciPy package (https://www.scipy.org/) in Python.

We investigated associations between the p(EC_50_) values and medical (available ICD-10 codes, active ingredients of prescribed medication) or demographic (sex, age) data, without binarization of p(EC_50_) values into hits and non-hits. The analysis included 588 ICD-10 codes assigned to at least 100 patients and 384 active ingredients (‘medications’) appearing at least 100 times, sex, and age. Situated in a Bayesian framework, we applied multivariable regression with logit-transformed EC_50_ values (i.e. -logit(EC_50_)) as outcome and with binary (0,1) predictor variables in the R package rstanarm (Goodrich et al., 2020), as shown before (Emmenegger, De Cecco, et al., 2023; Emmenegger, Emmenegger, et al., 2023; Emmenegger & Emmenegger, 2023), following the code set up by Julien Riou (Lamparter et al., 2022). Since regression using uninformative priors on regression coefficients (Normal(0,10)) led to the expected noise, given the number of parameters included in the analysis, we employed regularization techniques (Bayesian LASSO and regularized horseshoe priors, see (Gelman et al., 2013; Piironen & Vehtari, 2017)). We then depicted the associations between ICD-10 codes, medication, or demographic features and the respective -logit(EC_50_) values as odds ratio with 95% credible interval. Statistical testing of age in hits (‘seropositives) and non-hits (‘seronegatives’) was performed with Mood’s median test using the Python package scipy.stats.median_test (https://www.scipy.org/) or using Mann-Whitney U/Wilcoxon rank sum test at *α-level* 0.01. Statistical tests (and respective post-hoc corrections, e.g. when accounting for multiple comparisons) were always specified in the text and in the figure legend. To enhance visualisation, asterisks (*) were occasionally used in the figures. They denote significance in the following manner: p < 0.01: *, <0.001:**, <0.0001:***, <0.00001:****.

## Acknowledgments

We thank all the hospital patients and the healthy individuals included in this study for their contribution to this research project. We acknowledge the help of the Research Ward of the Clinical Trials Center of the University Hospital Zurich as well as of the Research Data Service Center of the USZ. The Yale Pediatric CF Center is acknowledged for the banking of serum samples, and Eugenie Chung for coordinating the shipping of biological material between institutions. We are also very grateful to all patients and families who consented to the study. ISAC 112 test for IgG4 RUO and IgE has kindly been performed by Service laboratory, Thermo Fisher Scientific, Alleröd, Denmark, Bjarne Kristensen. The library preparation for the *CFTR* sequencing using the FLUIDIGM kit was performed in collaboration with the Genetic Diversity Centre (GDC), ETH Zurich, with the great and kind help of Dr. Roberta Benedetto (Fluidigm AG). We thank the entire high-throughput screening team, University Hospital Zurich, for help with this project, namely Leyla Batkitar, Magdalena Bialkowska, Lisa Caflisch, Berre Doğançay, Julie Domange, Jingjing Guo, Marigona Imeri, Lidia Madrigal, Lorène Mottier, Rea Müller, Antonella Rosati, Dezirae Schneider, and Anne Wiedmer. Dr. Pål Johansen and Muriel Träxler (Dermatology, University Hospital Zurich) are acknowledged for help with ImmunoCAP, and Dr. Thomas M. Kündig for inspiring discussions. We thank Rita Moos from the University Hospital Zurich for help with IgG purification and Raphael Schoen (Zurich, Switzerland) for help with figure design. The team of Mabylon AG, Schlieren, particularly Natascha Wuillemin and Jürgen Pahla, have been helpful throughout these studies and have contributed with access to samples and knowledge – we are thankful for all the help! We acknowledge Dr. Jacobo Sarabia del Castillo and Dr. Stefano Vavassori (both Cellerys AG, Schlieren, Switzerland) for help with Fluorospot assays. Warm thanks to Dr. Cezmi Akdis and Dr. Mübeccel Akdis for hosting us at the SIAF in Davos and for all the enriching interactions with their group members. All authors wish to thank their groups and collaborators for support. Lastly, heartfelt thanks to a wonderful wife and most beloved friend and partner, Dr. Vishalini Emmenegger (formerly Bio Engineering Laboratory, ETH Zurich) for daily discussions, care, love, and for being infinitely inspiring beyond life and death.

## Author contributions

Study design and overall idea: AA, ME. Planned and conducted the high-throughput antibody profiling experiments shown in **Figs. 1-2** and **S1**: AA, ME. Analysed the data from the high-throughput antibody profiling experiments shown in **Figs. 1-2** and **S1**: ME, with contributions of AC. Analysed the adult CF data shown in **Figs. 3** and **S1** and performed the comparisons: ME. Conducted the antibody isotyping and IgG subclass experiments with ELISA and visualized the ImmunoCAP experiments shown in **Fig. 4A**-**D**: ME. Planned and conducted the competitive Western Blot experiments shown in **Fig. 4E**: AA, ME. Conducted, analysed, and visualized the Microfluidic Affinity Antibody Profiling experiments (**Fig. 4F**): MMS, GM, with help from TJPK. Carried out the ELISA-based experiments, analysed the data, and visualized the ISAC data shown in **Figs. 5** and **S2**: ME, with critical input from AA. Planned, conducted and analysed the Rapid extracellular antigen profiling data shown in **Figs. 6** and **S3**: YD, AMR, with analytical help from ME. Conducted the protein blast (not shown in a figure): ME. Conducted the BCR repertoire sequencing shown in **Figs. 7**, **S4**, **S5**, **S6**: CZ. Analysed the data from the BCR repertoire sequencing shown in **Figs. 7**, **S4**, **S5**, **S6**: FJN, HR, with help from CZ and ME. Planned, prepared, conducted, and analysed the HLA analysis (not shown in a figure): CZ, RM, MS, with help from ME. Planned, prepared, conducted, and analysed the T cell activation assays (**Fig. 7D**): RM, MS, ME. Had the idea to analyze *CFTR* in patients with allergies shown in **Fig. 8A-C**: AA. Prepared PCMBs and a DNA library of patients with peanut allergy for experiments shown in **Fig. 8A-C**: CZ, ME. Planned, prepared, and conducted the *CFTR* sequencing shown in **Fig. 8A-C**: ME, CZ. Analysed the data from the *CFTR* sequencing shown in **Fig. 8A-C**: NZ, with help from EMB and ME. Established a *CFTR* online questionnaire and analysed the data (**Fig. 8E-F**): ME, with help from KH and AA. Conducted a registry-based food allergy and *CFTR* study in Yale (not shown in a figure): SL, HN, with analytical help from ME. Planned, prepared, and conducted the peanut sensitisation in *Cftr* KO mouse model and related experiments (**Figs. 9 and S7**): EMB, LRH, RG, MEE, SCE, with analytical help from ME. Conducted and analysed the IFN-γ experiments shown in **Fig. S7D**: ME. Wrote the first version of the manuscript and conducted most analyses and created the figures: ME. Revised the manuscript and figures and suggested critical corrections: AA. Contributed to and critically revised the entire manuscript: all authors.

## Supplementary Figures

**Fig. S1. Additional illustrations pertaining to Fig. 1 and Fig. 2**.**A**. **A**. The average OD plotted against the respective p(EC_50_) values, for all samples with p(EC_50_) value ≥ 1.7. **B**. qq-plot showing distribution of p(EC_50_) values for nAra-h2 of all patient samples. **C**. Scatter dot plots visualizing PrP vs. nAra-h2 and CGRP vs. nAra h2. Histograms display the distribution of values. In blue colour: seropositives in the screen against nAra-h2, shown within the distribution of PrP and CGRP. **D**. Sample replicability in pwCF, some of which are seropositive and others seronegative.

**Fig. S2. ISAC data displaying raw values, pertaining to Fig. 5**. **A**. Raw values of the IgG4 assay are shown. **B.** Raw values of the IgE assay are shown.

**Fig. S3. Additional hit characterization of REAP experiment, pertaining to Fig. 6**. **A**. Number of hits, i.e. REAP score ≥ 2, for all conditions on the entire dataset. **B**. Number of hits, i.e. REAP score ≥ 2, for all conditions on the conformational proteins. **C**. Number of hits, i.e. REAP score ≥ 2, for all conditions on the protein epitopes. **A**-**C**. The red dotted line indicates the mean value. One-way ANOVA was performed to compare between all conditions. No statistically significant difference in the positivity rate was observed between either of the groups.

**Fig. S4. Additional illustrations pertaining to BCR repertoire sequencing shown in Fig. 7**. **A**. Number of sequences analysed. **B**. Sequences pertaining to respective immunoglobulin isotype, illustrated as fractions. Pairwise t-test indicated no significant differences between the groups. **C**. Distribution of heavy and light chain, illustrated as fractions. Pairwise t-test indicated no significant differences between the groups. **D**. VH family usage by isotype. Wilcoxon rank sum test was performed.

**Fig. S5. Additional illustrations pertaining to BCR repertoire sequencing shown in Fig. 7**. **A**. Evenness inferred from the Hill diversity index calculated over a range of diversity orders (q) ranging from 0 to 4, in equally spaced increments, for each subset. **B**. Mutation rate by VH for different isotypes. Wilcoxon rank sum test was performed.

**Fig. S6. Additional illustrations pertaining to BCR repertoire sequencing shown in Fig. 7**. No relevant convergence in sequences of known binders to peanuts were found using the data derived from BCR repertoire sequencing.

**Fig. S7. Additional illustrations pertaining to peanut sensitisation in mice shown in Fig. 9**. **A**. Shown are the distributions of the optical densities (ODs) irrespective of the plasma dilution after two weeks, five weeks, and post-challenge. **B**. Distributions of body temperature measurements post challenge among the different conditions. **C**. Same as in (B) but with anaphylactic score as readout instead of body temperature. Ps from Wilcoxon rank sum test with Holms correction for multiple comparisons. **D**. Determination of IFN- γ plasma concentrations in eight anti-Ara-h2-seropositive pwCF, three patients with systemic lupus erythematosus, 13 patients with mixed disease history, 10 COVID-19 patients, and six healthy controls. Distributional differences were assessed using Wilcoxon rank sum test, with CF as reference group for comparisons. No significant difference was observed before and after Holm’s correction for multiple comparisons. The assay was conducted in technical duplicates.

## Supplemental File

**Supplemental File 01**. Χ^2^ statistics for ICD-10 codes.

**Supplemental File 02**. Χ^2^ statistics for ICD-10 codes after removal of patients diagnosed with CF.

**Supplemental File 03**. Odds ratios for all ICD-10 codes that entered the Bayesian regression model.

**Supplemental File 04**. Χ^2^ statistics for active ingredients of medications.

**Supplemental File 05**. Odds ratios for all active ingredients of medications that entered the Bayesian regression model.

**Supplemental File 06**. Statistical comparisons of cohort screened for antibodies against multiple food proteins and controls.

**Supplemental File 07**. Data for all samples and proteins tested with ISAC.

**Supplemental File 08**. Data from BLAST experiment using BLOSUM-62 matrix.

**Supplemental File 09**. Data for all samples and proteins tested with REAP.

**Supplemental File 10**. HLA analysis.

**Supplemental File 11**. HLA sequence alignment.

**Supplemental File 12**. HLA binding prediction.

**Supplemental File 13**. Online survey codebook.

