## Supplementary figures and images for "The Cystic Fibrosis Transmembrane Regulator Controls Tolerogenic Responses to Food Allergens in Mice and Humans"

### Supplemental Figures

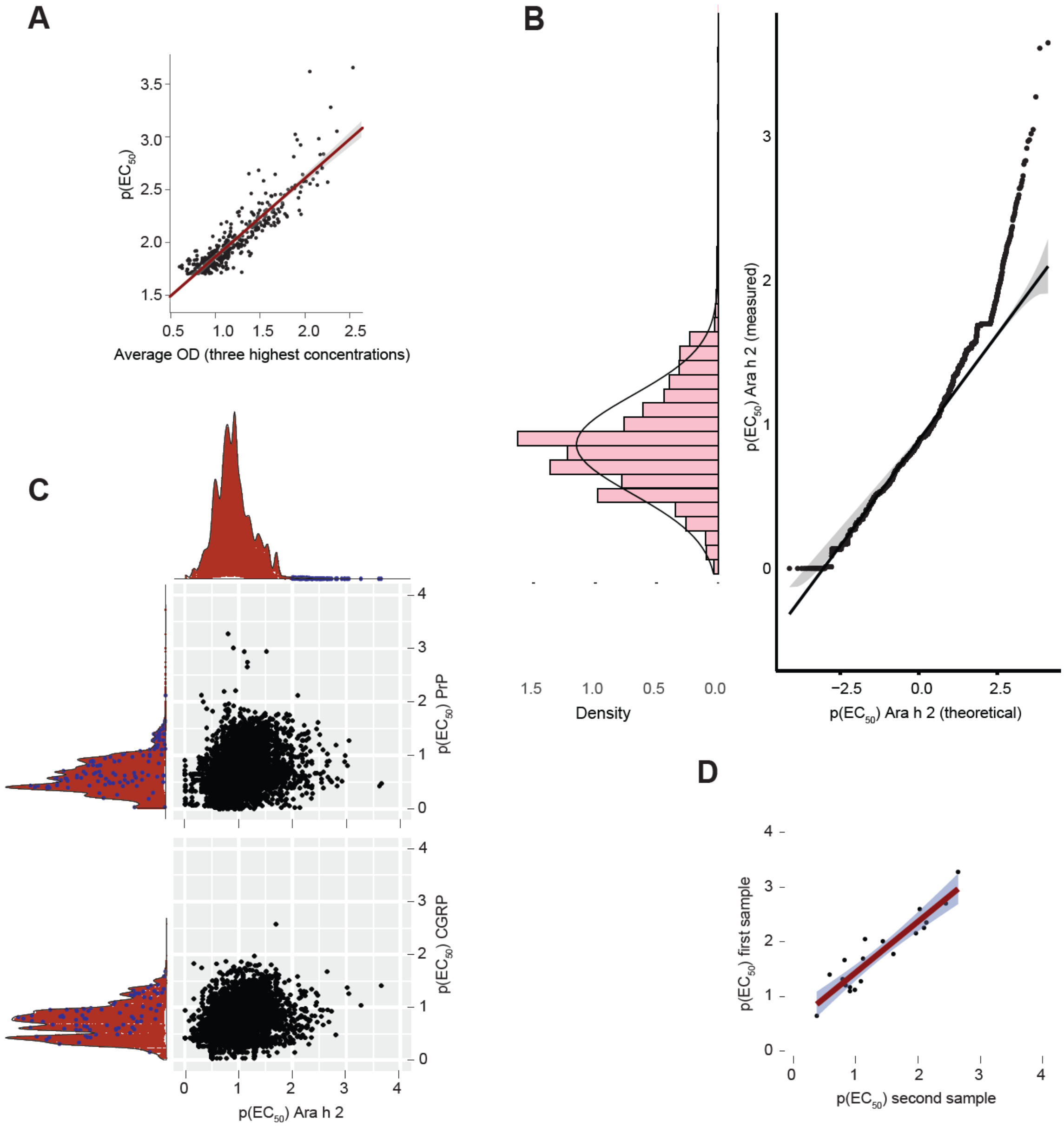

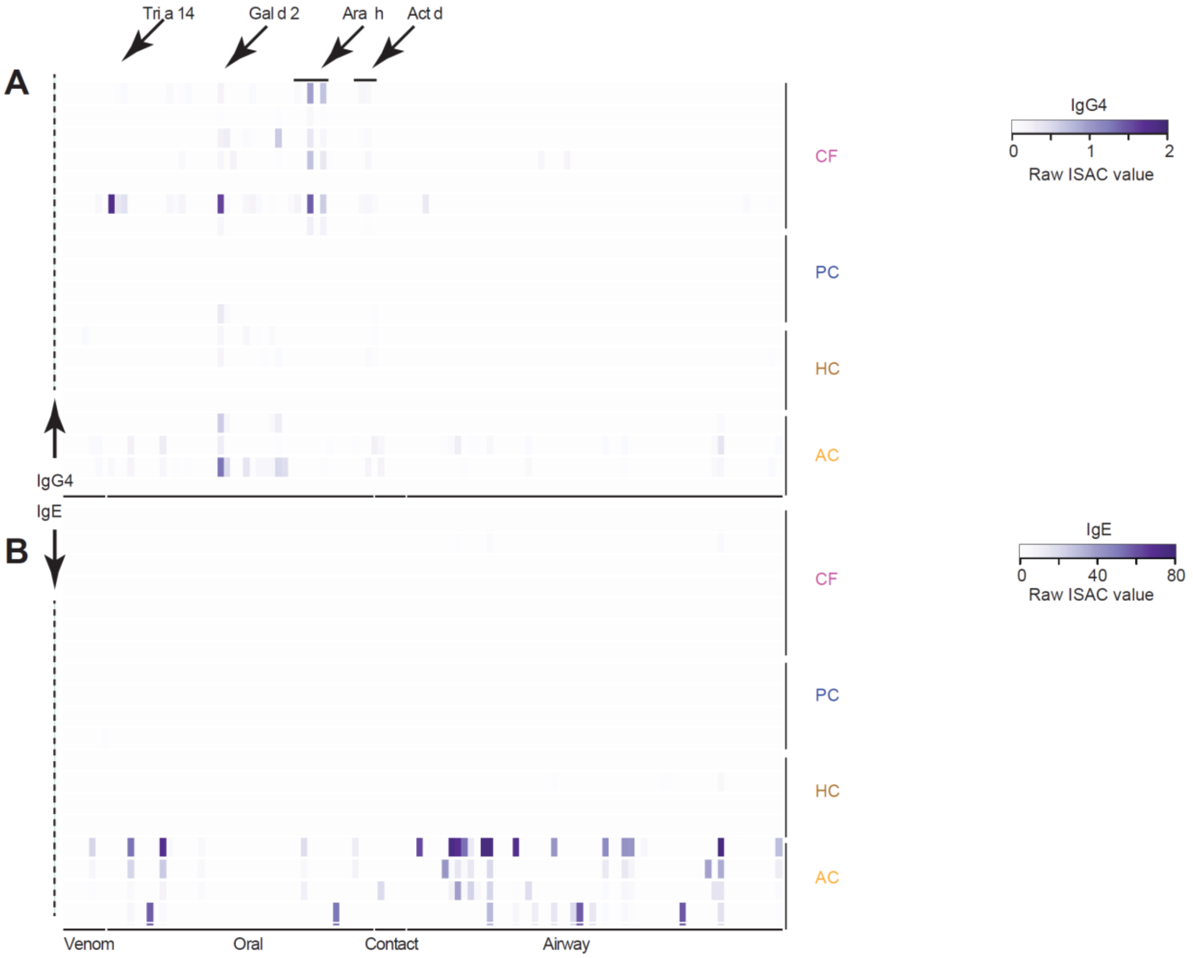

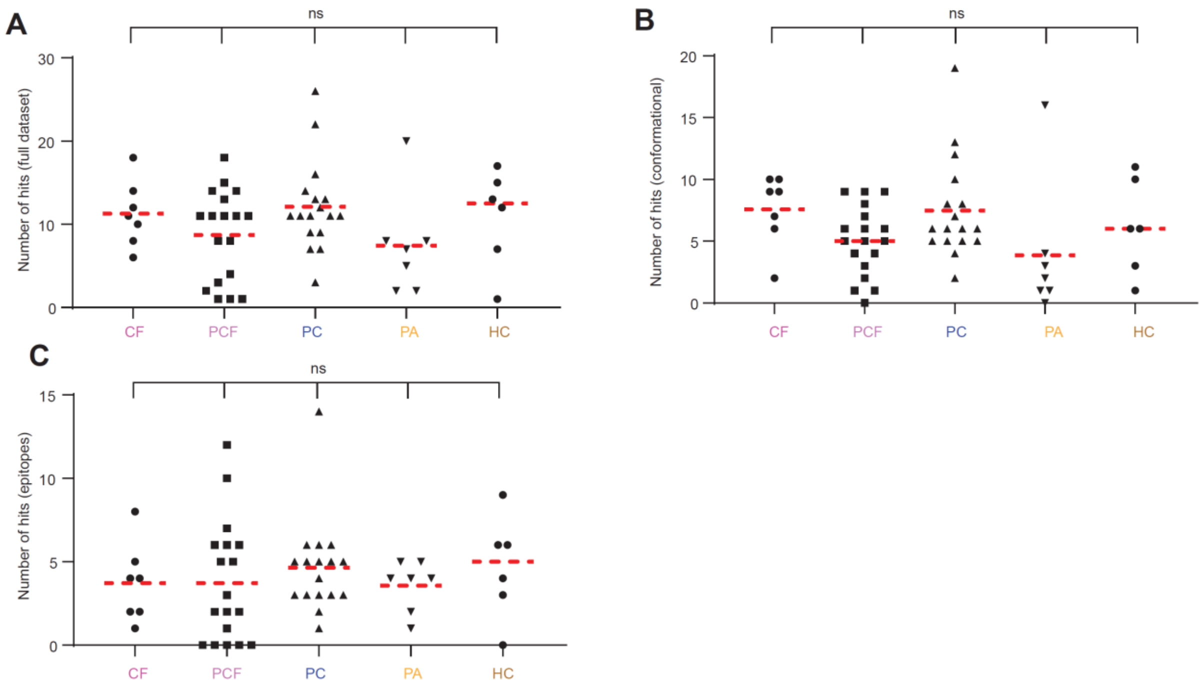

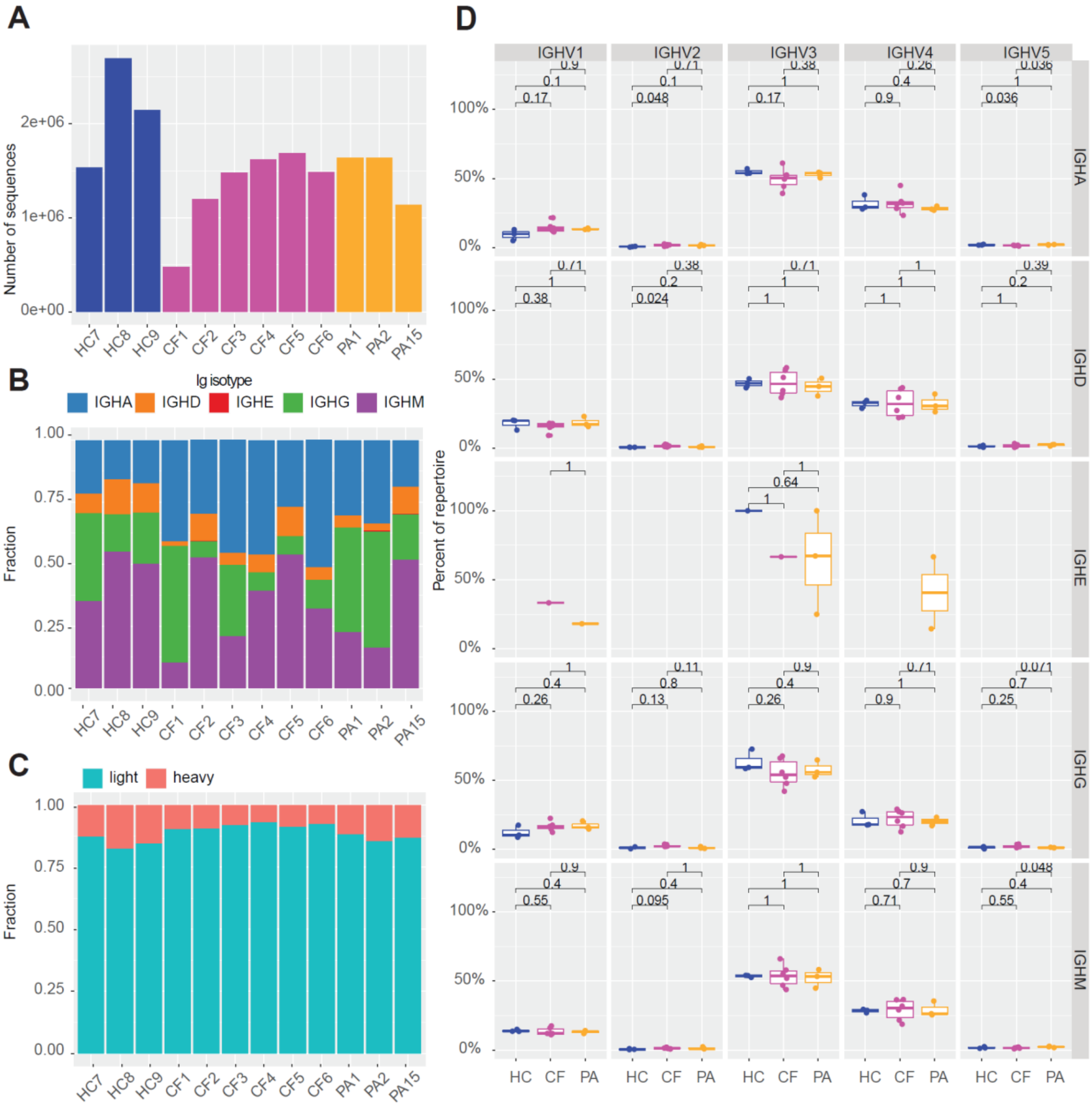

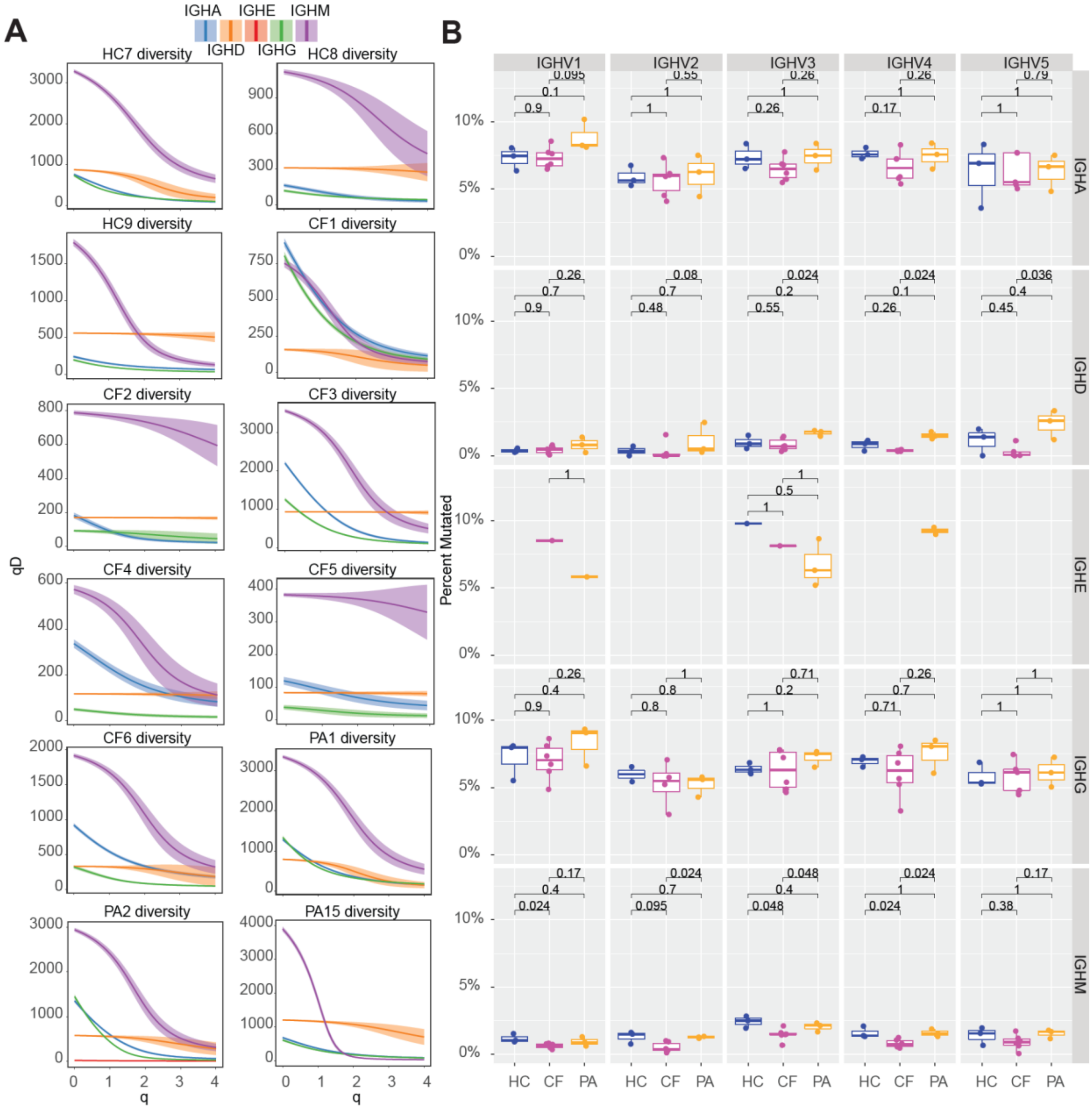

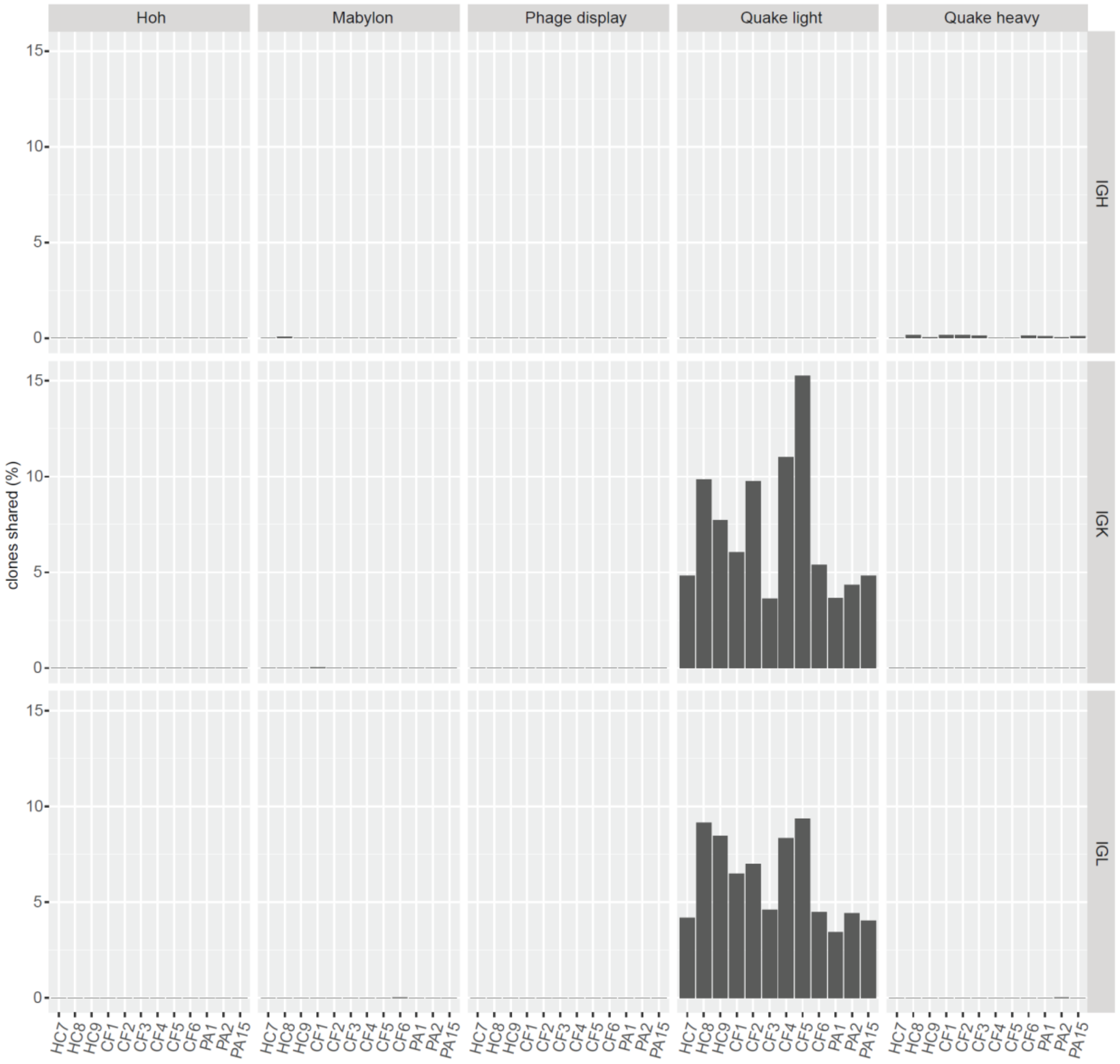

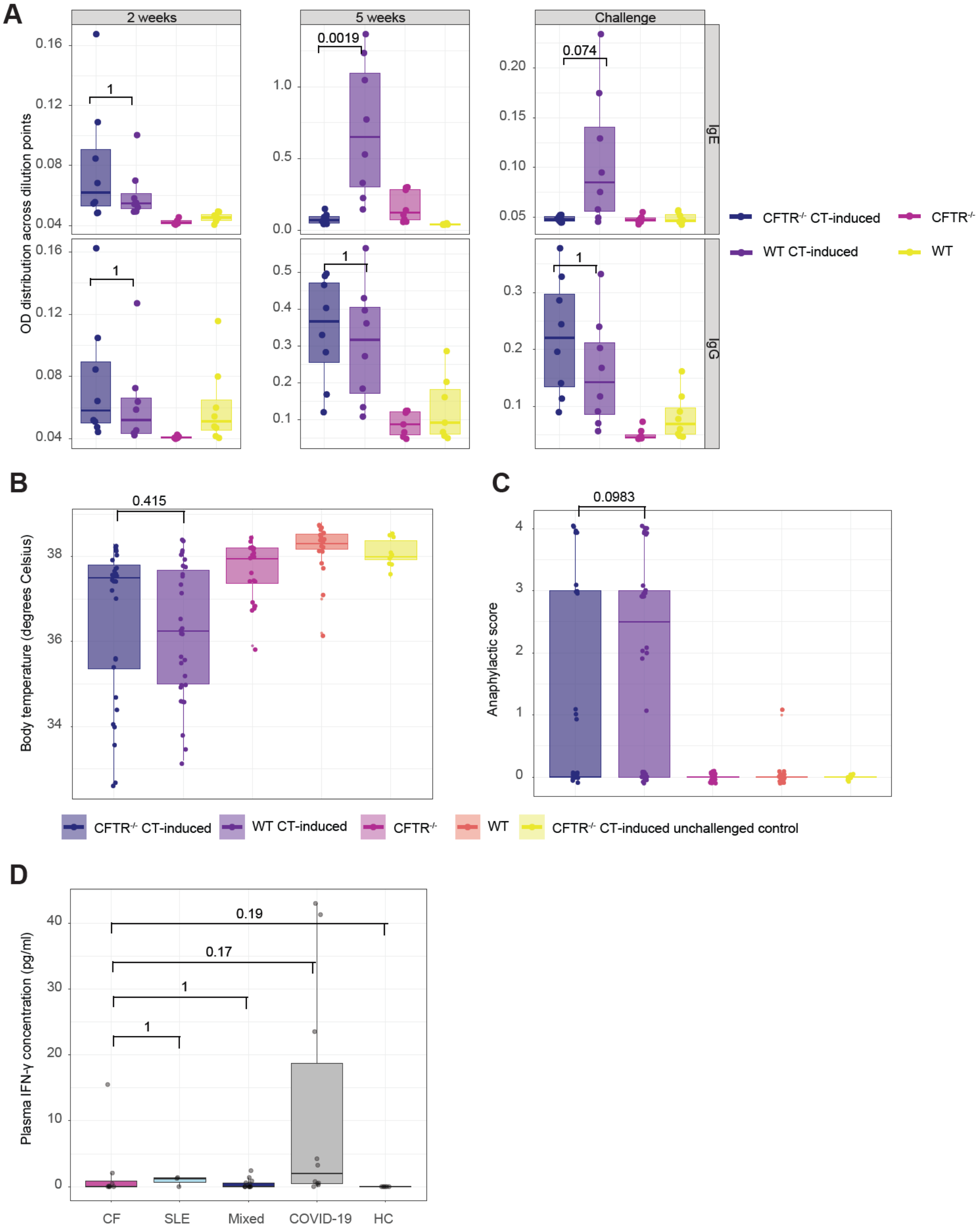
